# A Deep Learning Based Smartphone Application for Early Detection of Nasopharyngeal Carcinoma Using Endoscopic Images

**DOI:** 10.1101/2024.09.19.24313954

**Authors:** Yubiao Yue, Xinyu Zeng, Huanjie Lin, Jialong Xu, Fan Zhang, KeLin Zhou, Li Li, Zhenzhang Li

## Abstract

Nasal endoscopy is crucial for the early detection of nasopharyngeal carcinoma (NPC), but its accuracy relies significantly on the physician’s expertise, posing challenges for primary healthcare providers. Here, we retrospectively analysed 39,340 nasal endoscopic white-light images from three high-incidence NPC centres, utilising eight advanced deep learning models to develop an Internet-enabled smartphone application, “Nose-Keeper”, that can be used for early detection of NPC and five prevalent nasal conditions and assessment of healthy individuals. Our App demonstrated a remarkable overall accuracy of 92.27% (95% Confidence Interval (CI): 90.66%-93.61%). Notably, its sensitivity and specificity in NPC detection achieved 96.39% and 99.91%, respectively, outperforming nine experienced otolaryngologists. Explainable artificial intelligence was employed to highlight key lesion areas, improving Nose-Keeper’s decision-making accuracy and safety. Nose-Keeper can assist primary healthcare providers in diagnosing NPC and related diseases efficiently, offering a valuable resource for people in high-incidence NPC regions to manage nasal cavity health effectively.

## Introduction

Nasopharyngeal carcinoma (NPC) is the most common malignancy of the head and neck, particularly in East and Southeast Asia. Nonspecific early symptoms often lead to a delayed diagnosis, resulting in a suboptimal prognosis ^1–4^. The total survival rate of advanced cases over ten years typically falls between 50% and 70%. In contrast, the 5-year survival rate for promptly detecting nasopharyngeal carcinoma can approach 94%, highlighting the importance of early detection ^5–9^. Thus, devising a technique for timely NPC identification during clinical examinations is the primary aim of this study.

Nasal endoscopy plays a crucial role in the early detection of NPC ^5,10^. However, the accuracy of this examination relies heavily on the medical experience and expertise of the operators. Non-otolaryngology specialists, such as primary care doctors, emergency doctors, general practitioners, and paediatricians, may encounter difficulties interpreting endoscopic images owing to professional obstacles and inadequate expertise. They often overlooked the characteristic signs of NPC and confused them with those of nasal or nasopharyngeal diseases. This negligence frequently leads to missed diagnosis, misdiagnosis and delayed referrals, resulting in patients missing critical treatment windows ^11–13^. This issue is particularly pronounced in low- and middle-income countries where healthcare resources are limited and disease awareness is inadequate. Moreover, patients frequently overlook early nasopharyngeal cancer symptoms such as headaches and nasal congestion ^14,15^. Concurrently, financial constraints also lead to delayed medical consultations, increasing the risk of missing crucial early diagnosis and treatment ^9,16^. Consequently, to improve early detection rates and patient prognosis, it is essential to develop a novel, easy-to-use, and inexpensive method for early detection of NPC using endoscopic images.

Intelligent diagnostic solutions based on smartphones have enormous potential in the medical field, especially given the rapid growth of smartphone capabilities and the widespread application of deep learning algorithms ^17^. By analysing medical images immediately, these mobile health applications have demonstrated exceptional accuracy and efficiency in early disease identification and are becoming an emerging trend in healthcare ^18^. For example, advances in the early diagnosis of disorders such as keratitis, biliary atresia, ear infections, skin cancer, and lupus have been made, with some applications outperforming human expert performance ^19–24^. However, the creation of deep-learning smartphone applications for NPC remains an untapped research topic. Given the importance and complexity of identifying this malignancy, this gap highlights the importance and considerable potential of such applications. Therefore, exploring smartphone-based intelligent diagnostic methods for NPC promises to provide unique solutions for improving diagnostic accuracy and accessibility with significant scientific and practical significance.

In this study, we retrospectively collected 39,340 endoscopic white light images of 2,134 NPC patients and 11,824 non-NPC patients without NPC from three centres in high-incidence areas of NPC and developed eight advanced deep learning models with different architectures. Through validation, testing, and comparison with nine experienced otolaryngologists, we ultimately developed a smartphone application based on the Swin Transformer model called Nose-Keeper to improve the accuracy and efficiency of healthcare workers (especially primary healthcare providers) in diagnosing NPC, raise public’s awareness of NPC, and refer patients to professional medical institutions in a timely manner.

## Results

### Performance evaluation of various models in internal test set

**Table 1** presents the average overall accuracy, standard deviation, and corresponding 95% CI of the eight models for the internal dataset. The results indicate that the eight models developed achieved encouraging results in diagnosing seven types of nasal endoscopic images using transfer learning strategies and a large-scale dataset. The average overall accuracy of all the models exceeded 0.92. SwinT performed the best among the eight models, with an average overall accuracy of 0.9515. ResNet had the lowest average overall accuracy among the eight models, reaching 0.9221. From the standard deviation perspective, the most stable model was MaxViT, followed by SwinT. In addition, PoolF exhibited the highest standard deviation and the worst stability. In addition, **Supplementary Table 1** reports the time required for different models during the experimental process.

**Table 1.** Average overall accuracy of different models.

| Model | Fold1 | Fold2 | Fold3 | Fold4 | Fold5 | Average overall accuracy (95% CI) |
| --- | --- | --- | --- | --- | --- | --- |
| SwinT | 0.9519 | 0.9547 | 0.9519 | 0.9539 | 0.9449 | 0.9515 ($\pm 0.0039$) (0.9481-0.9549) |
| MaxViT | 0.9350 | 0.9448 | 0.9358 | 0.9456 | 0.9284 | 0.9379 ($\pm 0.0072$) (0.9316-0.9443) |
| PoolF | 0.9484 | 0.9531 | 0.9503 | 0.9523 | 0.9473 | 0.9503 ($\pm 0.0025$) (0.9481-0.9524) |
| CaiT | 0.9373 | 0.9448 | 0.9440 | 0.9480 | 0.9387 | 0.9426 ($\pm 0.0045$) (0.9387-0.9465) |
| ResNet | 0.9231 | 0.9369 | 0.9239 | 0.9342 | 0.9221 | 0.9280 ($\pm 0.0070$) (0.9219-0.9341) |
| DenseNet | 0.9393 | 0.9421 | 0.9385 | 0.9452 | 0.9331 | 0.9396($\pm 0.0045$) (0.9357-0.9436) |
| Xception | 0.9460 | 0.9460 | 0.9456 | 0.9476 | 0.9347 | 0.9440($\pm 0.0052$) (0.9394-0.9486) |
| ConvNeXt | 0.9409 | 0.9468 | 0.9495 | 0.9515 | 0.9390 | 0.9455($\pm 0.0054$) (0.9408-0.9503) |

**Table 2** reports the precision, sensitivity, specificity, and f1-score of eight models for diagnosing nasopharyngeal carcinoma. The experimental results showed that the developed models almost exceeded 0.9900 for all four indicators of nasopharyngeal carcinoma, except for the precision of ResNet. For sensitivity, SwinT achieved the best results, reaching 0.9984 (±0.0023) (0.9939-1.0000). For precision, specificity, and F1-score, PoolF achieved the best results, reaching 0.9959 (±0.0034) (0.9892-1.0000), 0.9992 (±0.0006) (0.9980-1.0000), and 0.9969 (±0.0012) (0.9945-0.9993), respectively. **Table 3** reports the performance of the eight models developed in diagnosing five non-NPC diseases and normal samples. Based on the results of evaluation metrics and the potential impact of model architecture on the performance of external testing, we chose SwinT, PoolF, Xception, and ConvNeXt as candidate models for the smartphone application.

**Table 2.** NPC Diagnosis performance (Average, standard deviation and 95% CI) of various models. ‘*’ represents the best result of the specific metric.

| Model | Precision | Sensitivity | Specificity | F1-score |
| --- | --- | --- | --- | --- |
| SwinT | 0.9949 ($\pm 0.0031$) | 0.9984 ($\pm 0.0023$) | 0.9991 ($\pm 0.0006$) | 0.9966 ($\pm 0.0015$) |
|  | (0.9888-1.0000) | (0.9939-1.0000)* | (0.9979-1.0000) | (0.9937-0.9995) |
| MaxViT | 0.9910 ($\pm 0.0145$) | 0.9938 ($\pm 0.0072$) | 0.9983 ($\pm 0.0027$) | 0.9923 ($\pm 0.0061$) |
|  | (0.9626-1.0000) | (0.9797-1.0000) | (0.9930-1.0000) | (0.9803-1.0000) |
| PoolF | 0.9959 ($\pm 0.0034$) | 0.9979 ($\pm 0.0022$) | 0.9992 ($\pm 0.0006$) | 0.9969 ($\pm 0.0012$) |
|  | (0.9892-1.0000)* | (0.9936-1.0000) | (0.9980-1.0000)* | (0.9945-0.9993)* |
| CaiT | 0.9949 ($\pm 0.0047$) | 0.9954 ($\pm 0.0033$) | 0.9991 ($\pm 0.0009$) | 0.9951 ($\pm 0.0011$) |
|  | (0.9857-1.0000) | (0.9889-1.0000) | (0.9973-1.0000) | (0.9929-0.9973) |
| ResNet | 0.9883 ($\pm 0.0066$) | 0.9964 ($\pm 0.0029$) | 0.9978 ($\pm 0.0012$) | 0.9923 ($\pm 0.0024$) |
|  | (0.9754-1.0000) | (0.9907-1.0000) | (0.9954-1.0000) | (0.9876-0.9970) |
| DenseNet | 0.9918 ($\pm 0.0033$) | 0.9964 ($\pm 0.0043$) | 0.9985 ($\pm 0.0006$) | 0.9941 ($\pm 0.0027$) |
|  | (0.9853-0.9983) | (0.9880-1.0000) | (0.9973-0.9997) | (0.9888-0.9994) |
| Xception | 0.9949 ($\pm 0.0018$) | 0.9969 ($\pm 0.0034$) | 0.9991 ($\pm 0.0003$) | 0.9958 ($\pm 0.0021$) |
|  | (0.9914-0.9984) | (0.9902-1.0000) | (0.9985-0.9997) | (0.9917-0.9999) |
| ConvNeXt | 0.9933 ($\pm 0.0061$) | 0.9948 ($\pm 0.0075$) | 0.9988 ($\pm 0.0011$) | 0.9940 ($\pm 0.0037$) |
|  | (0.9813-1.0000) | (0.9801-1.0000) | (0.9966-1.0000) | (0.9867-1.0000) |

**Table 3.**
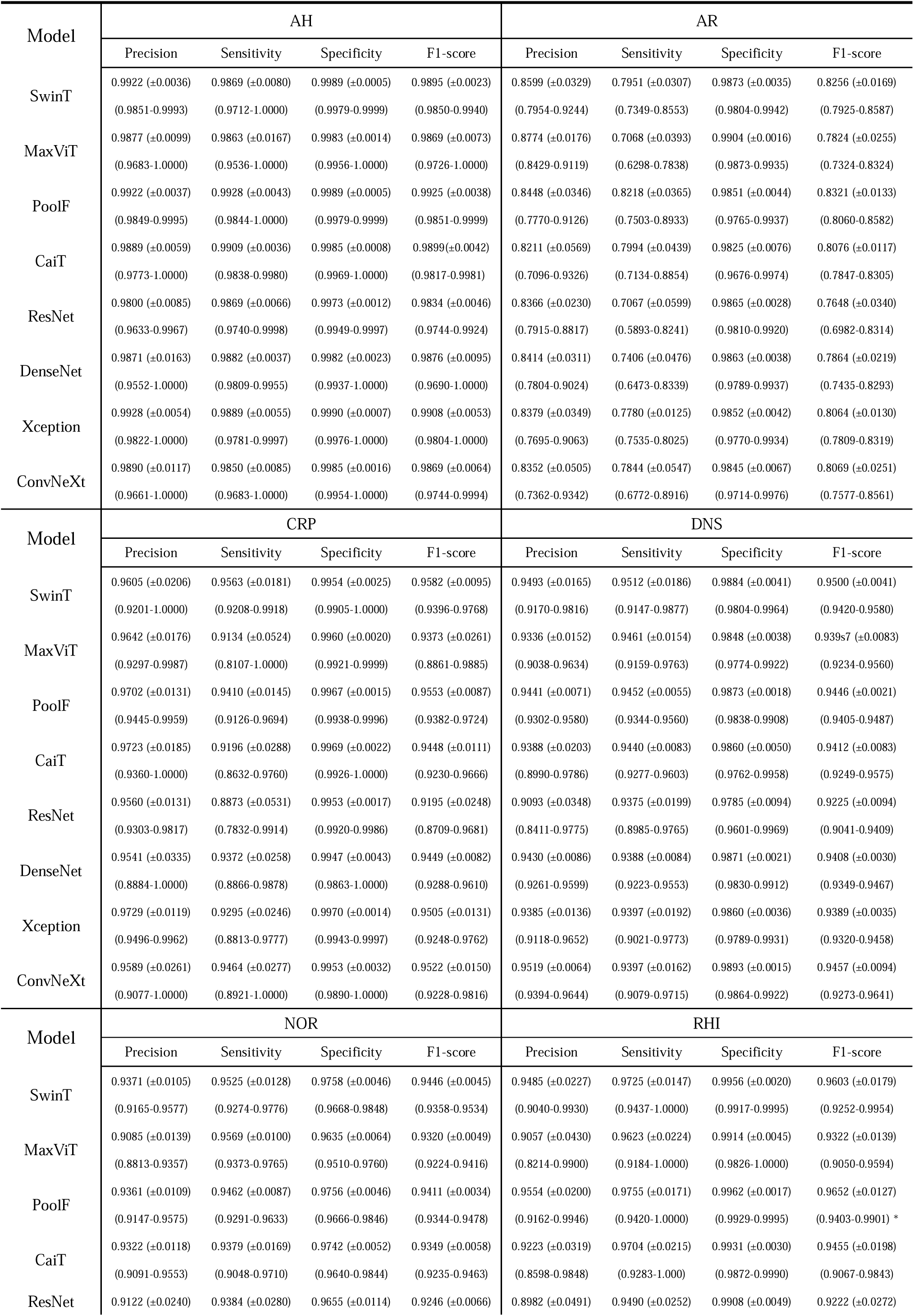
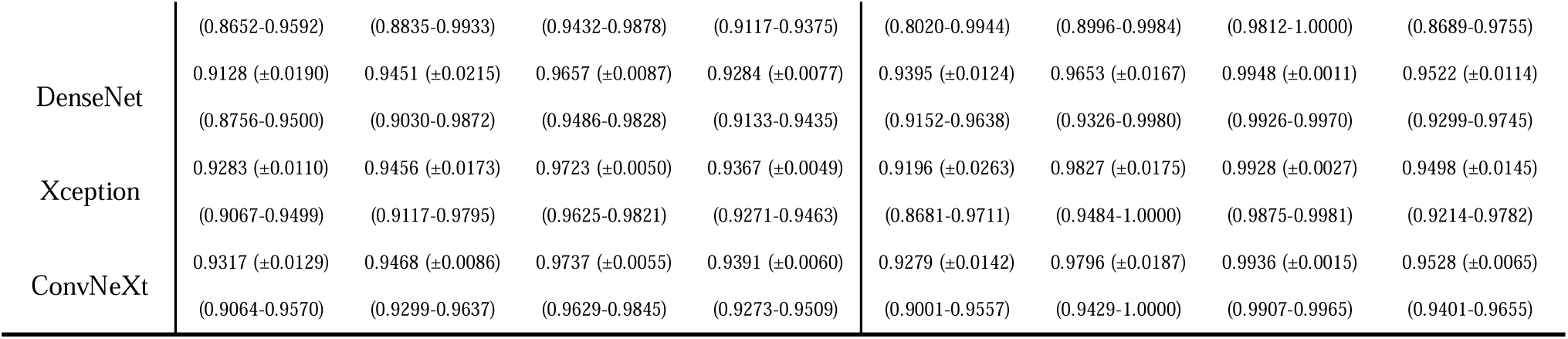
Non-NPC diagnosis performance (Average, standard deviation and 95% CI) of various models.

We initialised each candidate model parameter using the best weight from the five-fold cross-validation and then used the corresponding internal validation set to determine the optimal temperature for each candidate model when using a temperature scaling strategy. **Figure 1** shows the result changes of calibration metrics (Brier-score and Log-Loss) for each model on the internal test set. The experimental results indicate that the pre- and post-calibration results (**Figure 1a-b**) of SwinT were the best among the candidate models. In comparison, the results of the remaining candidate models were obviously inferior to SwinT (**Figure 1c-h**).

**Figure 1:**
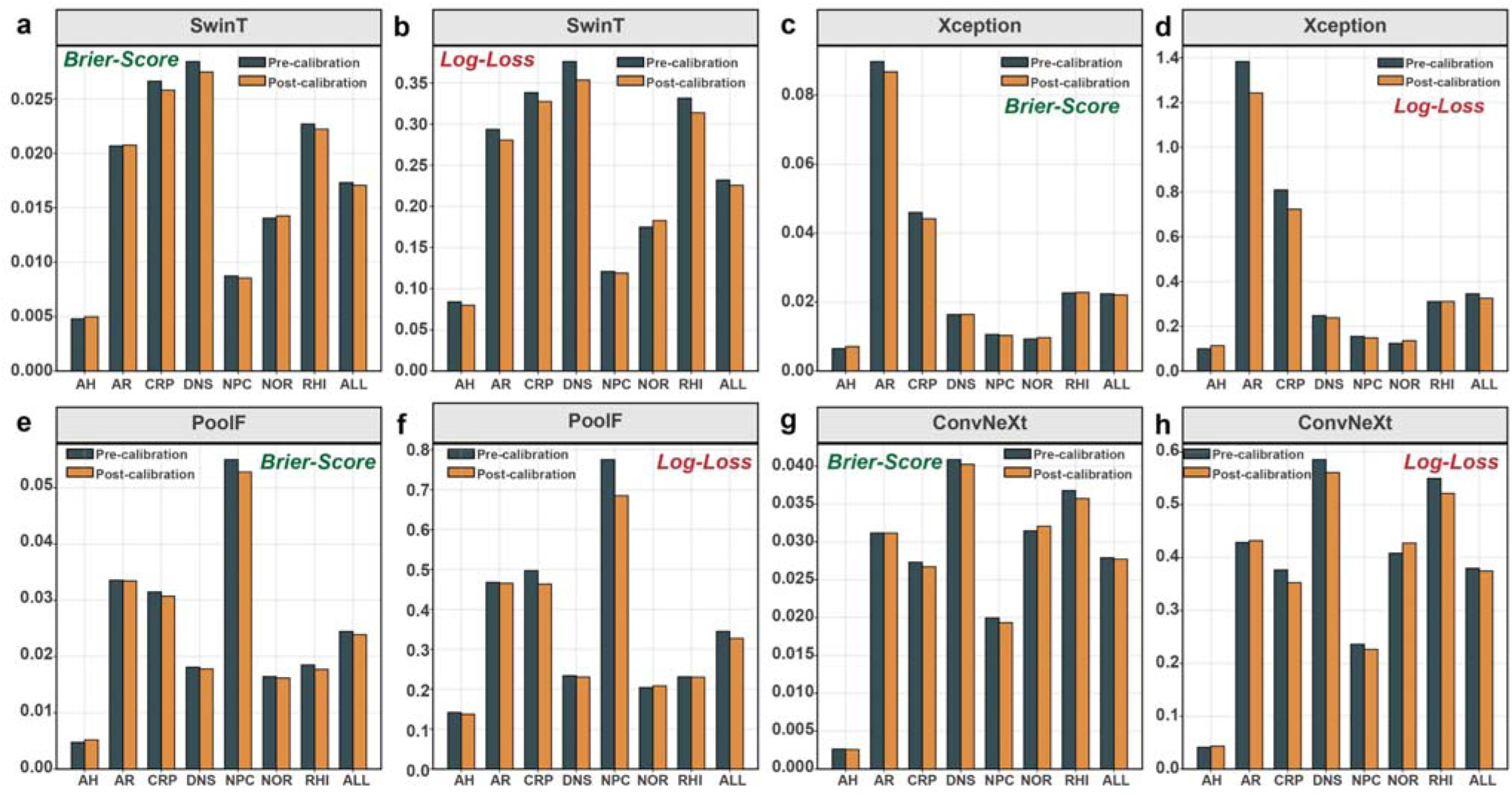
The result changes of calibration metrics of four candidate models in diagnosing different categories. The result changes of Brier-Score and Log-Loss of each model for each category were plotted. **a** Brier-Score of SwinT. **b** Log-Loss of SwinT. **c** Brier-Score of Xception. **d** Log-Loss of Xception. **e** Brier-Score of PoolF. **f** Log-Loss of PoolF. **g** Brier-Score of ConvNeXt. **h** Log-Loss of ConvNeXt. Particularly, “**ALL**” in X-axis means the calibration performances of the entire external test set.

### Testing the candidate models in external test set

Four candidate models were tested using the external test set from LZH to further evaluate their performances in real-world clinical settings. **Figure 2** shows the overall accuracy and confusion matrix of the four candidate models on the external test set. All the predicted results were shown after calibration. The confusion matrix was used to analyse the sensitivity and specificity of each model for a specific category. The experimental results indicate that SwinT (**Figure 2a**) achieved a state-of-the-art performance, far superior to Xception (**Figure 2b**), PoolF (**Figure 2c**) and ConvNeXt (**Figure 2d**). Concretely, it achieved the highest overall accuracy, reaching 92.27% (95% CI: 90.66%-93.61%). For NPC, the sensitivity and specificity of SwinT reached 96.39% (95% CI: 92.74%-98.24%) and 99.91% (95% CI: 99.47%-99.98%), respectively (**Figure 2a**). For non-NPC categories, the sensitivity and specificity of the SwinT exceeded 86.00% and 95.00%, respectively. **Figure 3** shows the ROC curves of the four candidate models. In terms of the ROC curve, the SwinT was also found to be the best model (**Figure 3a**). SwinT’s AUCs for all seven categories were greater than 0.9900. For NPC, SwinT’s AUC reached 0.9999 (95% CI: 0.9996-1.0000). In contrast, the AUCs of Xception (**Figure 3b**), PoolF (**Figure 3c**), and ConvNeXt (**Figure 3d**) for NPC were 0.9994 (95% CI: 0.9985-0.9999), 0.9916 (95% CI: 0.9856-0.9958), and 0.9989 (95% CI: 0.9976-0.9997), respectively,

**Figure 2:**
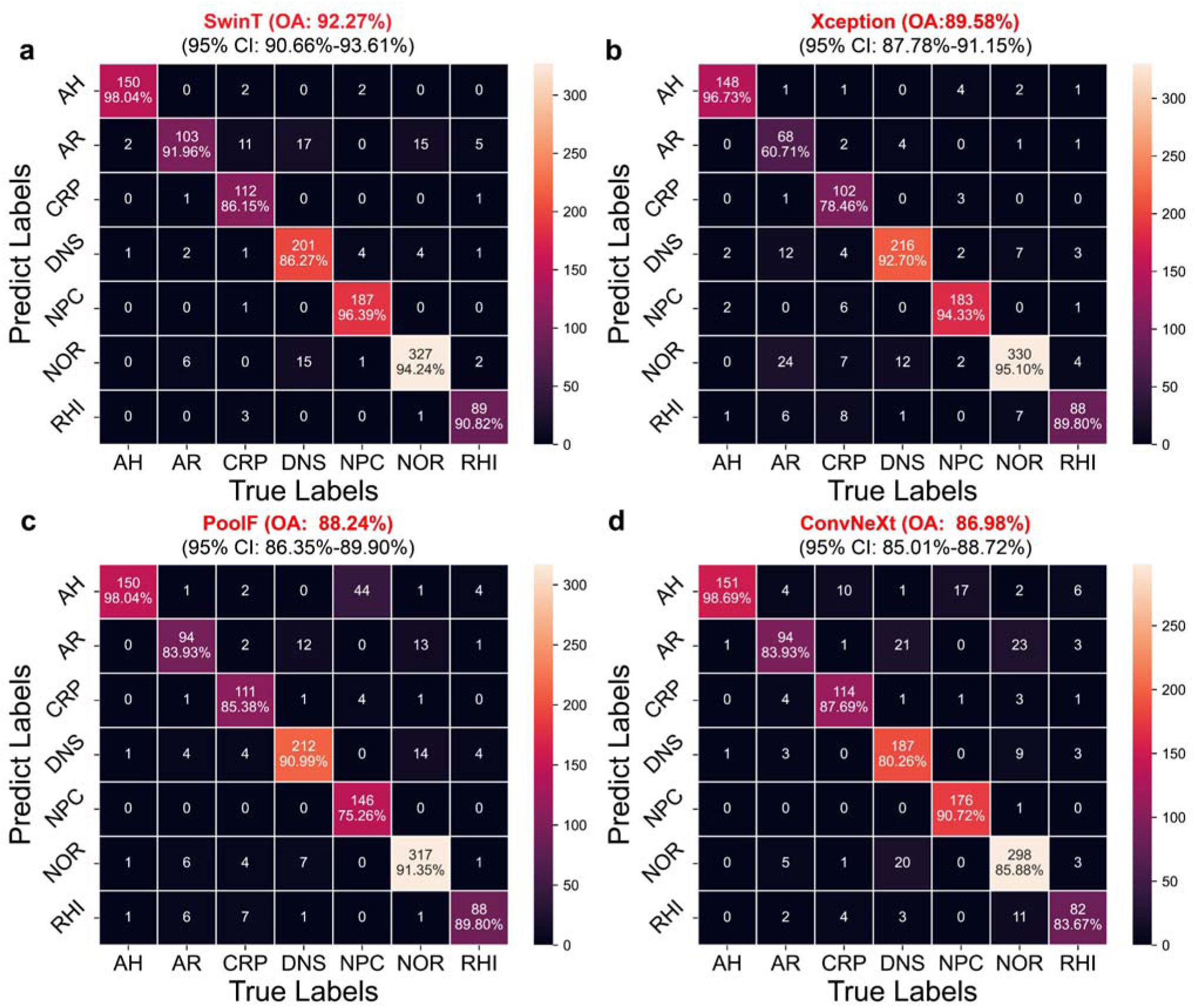
Confusion matrix of four candidate models in the images from LZH. The figure also reports the overall accuracy of each model on the external test set and the corresponding confidence interval. **a** The confusion matrix of the SwinT. **b** The confusion matrix of the Xception. **c** The confusion matrix of the PoolF. **d** The confusion matrix of the ConvNeXt.

**Figure 3:**
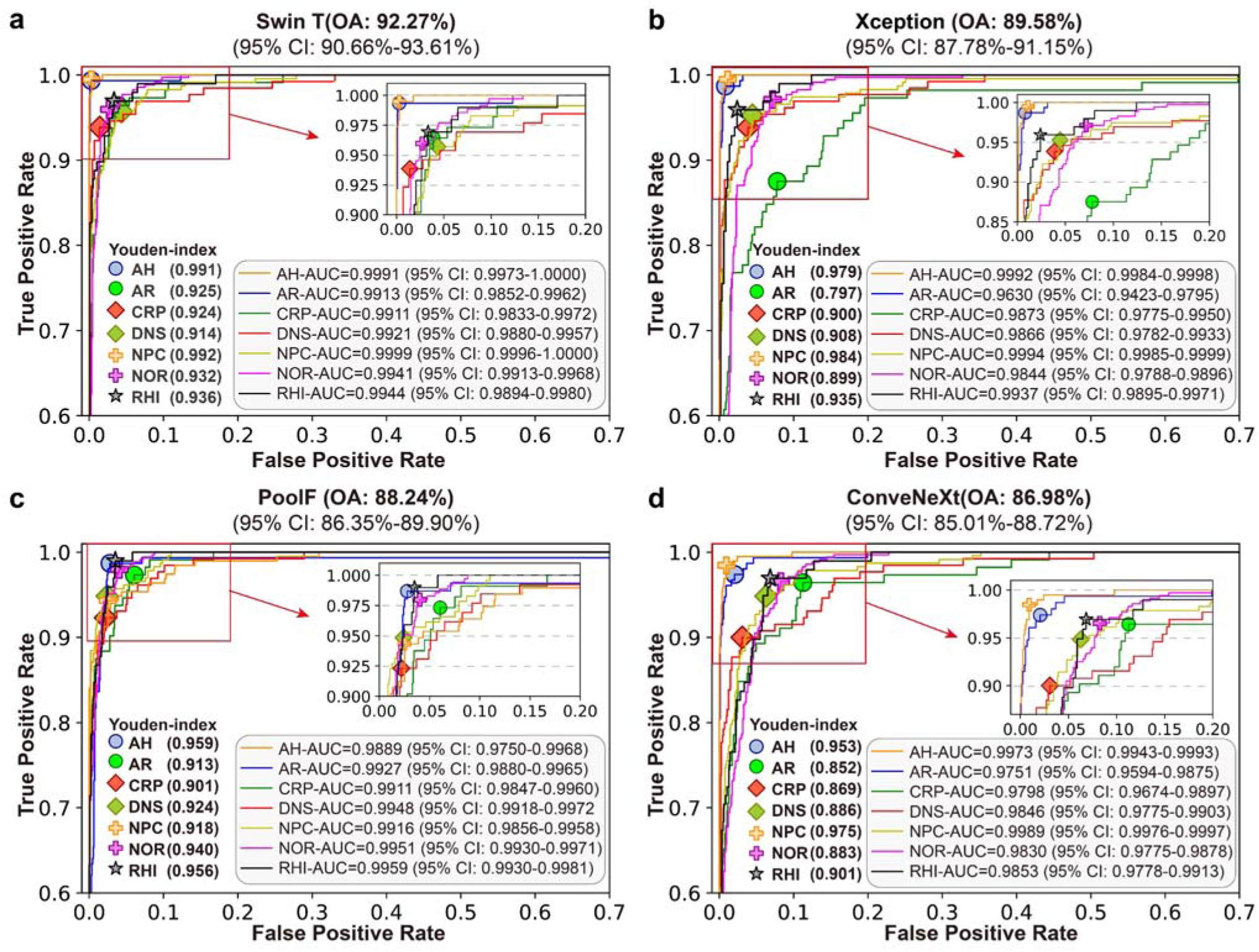
Receiver operating characteristic curve (ROC) and optimum Youden-index results of candidate models. **a** The ROC and optimum Youden-index results of SwinT. **b** The ROC and optimum Youden-index results of Xception. **c** The ROC and optimum Youden-index results of PoolF. **d** The ROC and optimum Youden-index results of ConvNeXt.

However, compared to the internal test set, the performances of the four candidate models decreased on the external test set. We initially extrapolated that this phenomenon might be caused by the different imaging equipment and image acquisition process in the external test set. **Figure 3** also presents the optimum Youden index results for each model for the different categories to compare the performance of the different models further. Experimental verification showed that SwinT was the best model for diagnosing NPC, with a Youden index of 0.992. SwinT also demonstrated excellent performance in diagnosing AH, AR, and CRP with Youden indices of 0.991, 0.925, and 0.924, respectively. The results fully demonstrated the application potential of SwinT in diagnosing different categories. Overall, the experimental results for the external test set indicated that SwinT performed the best among the four candidate models. Therefore, SwinT was chosen to deploy the smartphone application.

### Robustness of the SwinT in the external test set

**Figure 4** reports the results of the robustness analysis of the SwinT. **Figure 4a** illustrates examples from the external test set using various transformation strategies. **Figure 4b-f** details the performance metrics (overall accuracy, sensitivity, precision, specificity, and f1-score) of SwinT across the 12 enhanced datasets. The SwinT showed good robustness to rotation changes, likely benefiting from the training of the model with random rotation augmentations. For Gaussian blur transformation, the performance of the model decreased with an increase in the blur level, but the overall performance remained relatively stable. When using a slight decrease (Brightness I) or increase (Brightness II) in brightness, the accuracy of the model was minimally affected by interference, and its overall performance remained relatively steady. However, an excessive increase in brightness caused a significant decrease in model performance, affecting the sensitivity (AH, CRP, NPC, and RHI), precision (AH), F1-score (AH, CRP, and NPC), and specificity for AH. Similarly, when applying slight decreases (Saturation I) and increases (Saturation II) in the saturation, the accuracy of the model was not significantly affected. However, significant saturation enhancement (Saturation III) led to notable deviations in accuracy, particularly affecting the sensitivity for AH, precision for CRP and RHI, F1-score for AH and CRP, and specificity for CRP and RHI.

**Figure 4:**
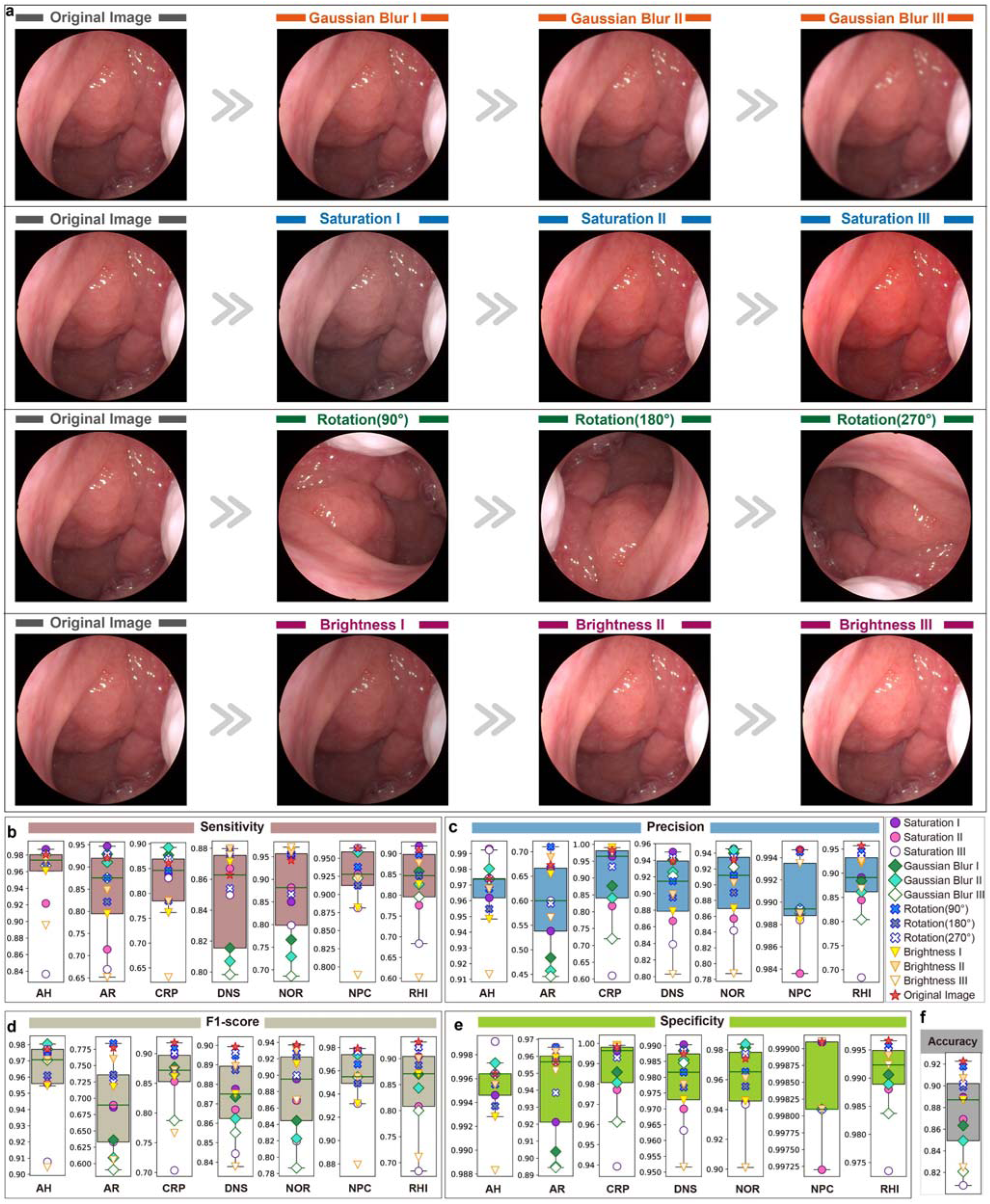
Performance comparison of SwinT on the external test dataset using 12 image transformations. **a** Examples of 12 image transformations. **b** Sensitivity of SwinT for external test set. **c** Precision of SwinT for external test set. **d** F1-score of SwinT for external test set. **e** Specificity of SwinT for external test set. **f** Overall accuracy of SwinT for external test set.

### Heatmap and Comparison of diagnostic results between SwinT and otolaryngologists

**Figure 5** visually explains the model’s internal decision-making mechanism and represents the results of the human-machine comparison experiment. **Figure 5a** shows the heat maps generated by SwinT using the Grad-CAM algorithm for seven types of nasal endoscopic images. Experimental results indicated that SwinT could effectively focus on the key areas of each type of image. Visually, the colour distribution of the heat map conformed to the professional insights of otolaryngologists. For NPC, Grad-CAM effectively helped SwinT to highlight the lesion area.

**Figure 5:**
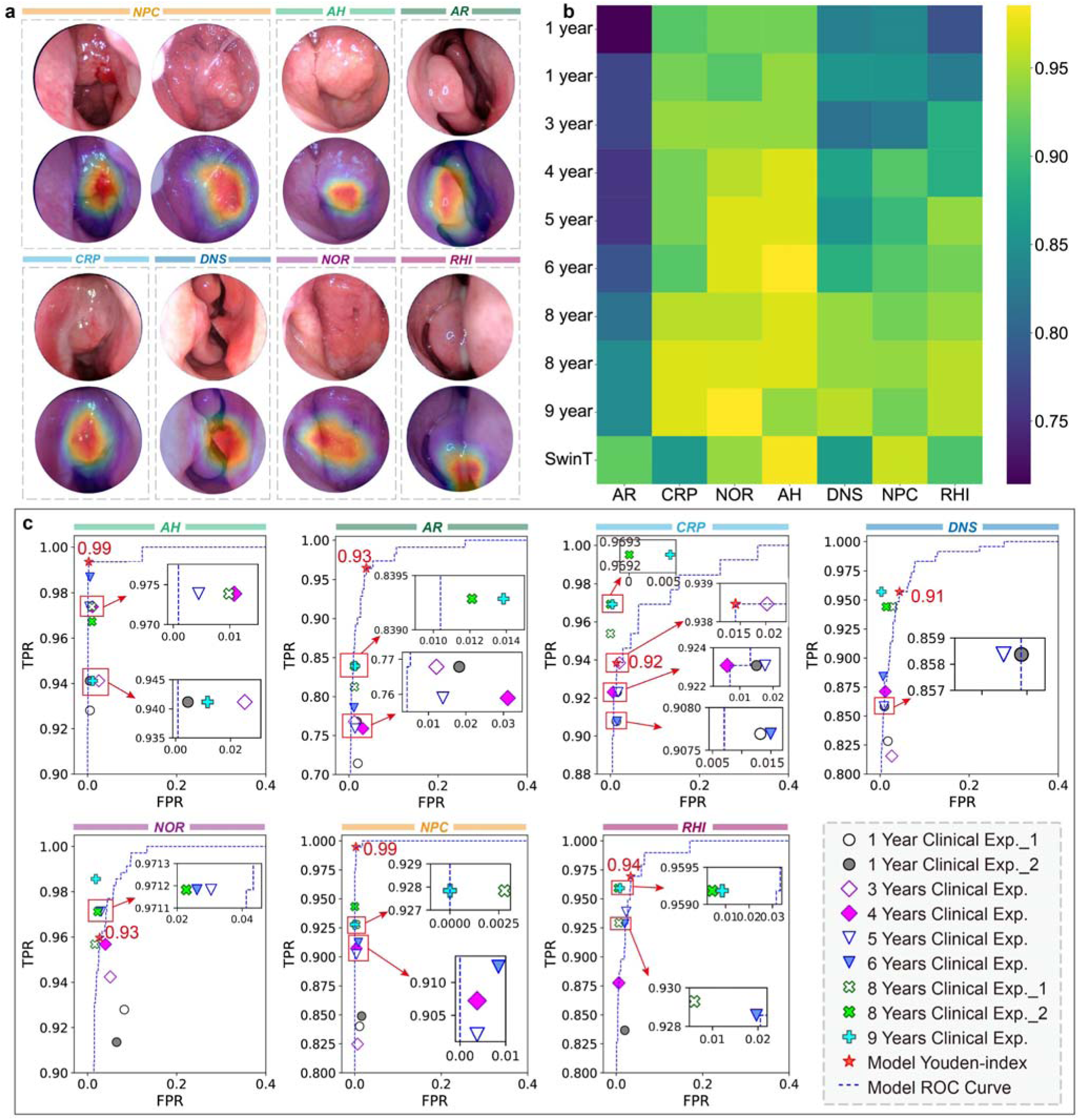
The heatmaps and Human-machine comparison results. **a** the heatmaps of different nasal endoscopic images generated by Grad-CAM. **b** Comparison of the sensitivity between SwinT and otolaryngologists. **c** The ROC curve of SwinT, the optimum Youden-index results of SwinT and the otolaryngologists.

**Figure 5b** shows the performance of SwinT and the nine otolaryngologists in diagnosing different diseases. In **Figure 5b**, the closer the color was to green or yellow, the more accurate the doctor or model was in diagnosing the disease. The average sensitivity of the nine physicians for NPC was 0.8927. Among them, the best doctor to diagnose NPC was a doctor with eight years of clinical experience, with a sensitivity of 0.9433, and the worst doctor was a doctor with three years of clinical experience, with a sensitivity of 0.8247. Obviously, SwinT outperformed all experts in diagnosing NPC. Furthermore, for AR, SwinT outperformed all experts. Because SwinT was prone to misjudge CRP as an AH, its performance was not as good as that of experts. For AH, the SwinT was superior to most other physicians. The SwinT was superior to some doctors in terms of the DNS, NOR, and RHI scores. **Figure 5c** and **Supplementary Figure 1** show the performance differences between SwinT and the nine otolaryngologists in diagnosing various types of endoscopic images in more detail. Based on the optimum Youden index results of SwinT and otolaryngologists, we concluded that SwinT outperformed all otolaryngologists in diagnosing AH, AR, NPC, and RHI. When diagnosing the remaining three types of endoscopic images, SwinT was slightly inferior to some otolaryngologists (clinical experience: from five years to nine years).

### Display of the Smartphone Application

The main function of Nose-Keeper (**Figure 6**) is to read real-time images captured by a nasal endoscope connected to an Android phone with a Micro USB interface or to load local nasal endoscope images (for example, the user can capture images from other endoscope devices, and then the image is uploaded to the smartphone album) (**Figure 6a**). Subsequently, by clicking the one-click detection button (**Figure 6b**) on the application, the user can obtain the diagnosis results of the image using the AI model. In addition, we listed a heat map corresponding to the original image on the results page to enhance the security of the application and to remind users to pay attention to the diseased area actively (**Figure 6c**). Nose-Keeper can also read multiple endoscopic images simultaneously and use a voting mechanism to further improve prediction accuracy (**Supplementary Note 6**). In particular, we listed reference images of various diseases and some common medical sense to improve the users’ understanding of diseases and medical procedures. Such an intelligent application will potentially help the majority of primary care providers who lack clinical experience and professional knowledge in diagnosing nasal diseases, and the public residing in high-risk areas will primarily judge whether the captured nasal endoscopic image contains NPC, common nasal cavities, and nasopharynx diseases. We tested the running speed of Nose-Keeper using four different Android smartphones (i.e., Xiaomi 14, Xiaomi 12S Pro, HUAWEI nova 12, and HUAWEI mate 60) at a network speed of 100Mb/S. The results show that the time consumption is approximately 0.5s to 1.1s. For more detailed feature introductions and user pages of Nose-Keeper, please refer to **Supplementary Figure 2** and **Supplementary Movie 1**.

**Figure 6:**
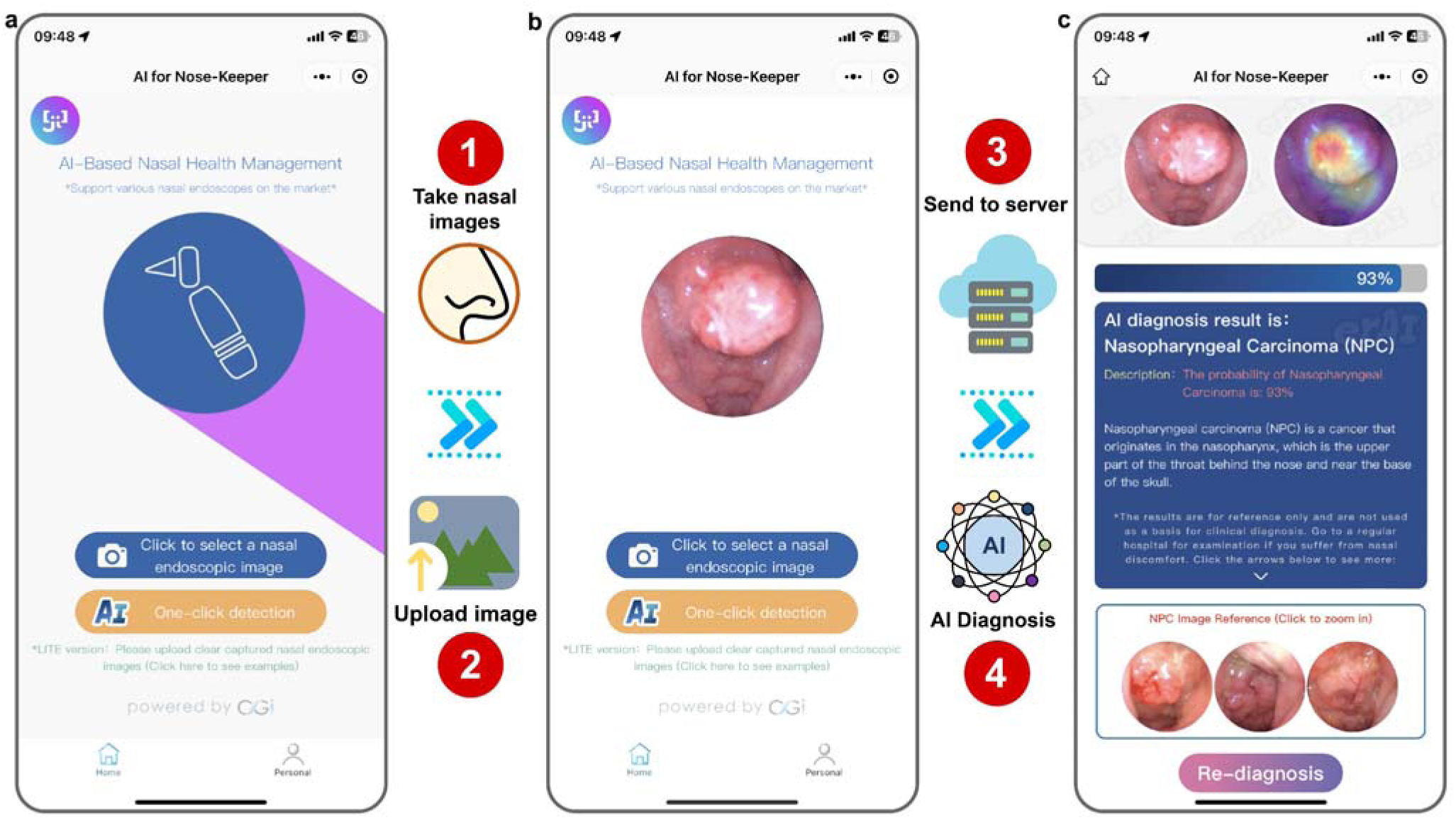
The application page and usage process of Nose-Keeper. The application process mainly includes login, upload image, submit identification, and finally output detection result report. **a** The home page after login. **b** The page after uploading the image. **c** The result page after data processing.

## Discussion

To the best of our knowledge, this study is the first to develop a smartphone application based on a deep learning model named Nose-Keeper to diagnose NPC and non-NPC effectively. To ensure the practicality of Nose-Keeper, we retrospectively collected 6,014 NPC white-light endoscopic images and 33,326 white-light endoscopic images of common diseases of the nasal cavity, nasopharynx, and normal nasal cavities from three hospitals and trained eight different deep learning models. In this study, to shorten the training time of the model and improve the generalisation of the model as much as possible, we used a popular transfer learning strategy. Through extensive evaluation and testing (including model metric comparison, model calibration, robustness analysis, and human-machine comparison), we found that the developed SwinT reached state-of-the-art application potential and encapsulated it into Nose-Keeper. Compared to nine otolaryngologists with different diagnostic experiences, Nose-Keeper outperformed all otolaryngologists in diagnosing NPC. For diseases other than NPC, the diagnostic performance of the model was comparable to that of most physicians. In addition, we used the Grad-CAM algorithm in the application to visually display the areas in the image that affect the decision-making results of the model, which effectively reminds users to pay attention to the lesion area. Because the Nose-Keeper is deployed on a cloud server, its operation is not affected by the hardware. Users only need to use the Internet to obtain diagnostic results from the Nose-Keeper in real-time.

Previously, researchers in the field of computer vision used convolutional neural networks to process various images. Researchers have recently begun to focus on powerful architectures of Vision Transformers. Initially, Vision Transformers were applied to natural language processing systems such as the recently popular large language models. Compared with traditional convolutional neural networks, Vision Transformers rely on a self-attention mechanism to achieve better results in image processing. Considering that Vision Transformers are attracting more and more attention and may be more suitable for processing medical images, we tentatively trained four Vision Transformers and four convolutional neural networks in this work to obtain the best model. The experimental results demonstrated that most of the selected Vision Transformers were better than convolutional neural networks for diagnosing nasal endoscopic images. In particular, we found that Swin Transformer achieved state-of-the-art performance on both the internal and external test sets. From the perspective of Swin Transformer’s internal modeling mechanism, Swin Transformer is essentially a hierarchical Transformer that uses shifted windows. Swin Transformer constructs a hierarchical representation by starting from small-sized patches and gradually merging neighboring patches in deeper Transformer layers ^25^. By employing the shifted window based self-attention, Swin Transformer only calculates self-attention within a local window, which greatly reduces the computational complexity. Meanwhile, in consecutive Swin Transformer blocks, the shifted windowing scheme allows for cross-window connection, i.e., the model can shift the windows in a certain pattern to ensure that the lesion feature information flows between different windows. The unique mechanism of the Swin Transformer enables it to maintain the advantages of the Vision Transformers in modeling long-range dependencies while effectively capturing local information in nasal endoscopy images, thereby improving the accuracy in diagnosing NPC and non-NPC diseases. These findings provide practical guidance for nasal endoscopy researchers.

The World Health Organization (WHO) Global Observatory for eHealth (GOe) defines mHealth as medical and public health practice supported by mobile devices ^26^. mHealth has the potential to change healthcare and support public health and primary healthcare ^27^. With the booming development of smartphones in our daily lives, the combination of advanced medical technology and mHealth to manage diseases has become an unstoppable trend ^28^. Nowadays, smartphones have become an indispensable part of daily life, and mHealth applications have found a place in healthcare systems ^29^. On a global scale, the penetration rate of smartphones reached 68% in 2022 ^30^. Especially in developing countries, the number of smartphone owners is constantly increasing, leading to significant social and economic changes ^31^. In 2022, nearly nine in 10 internet users in Southeast Asia located in the high incidence area of NPC will use smartphones this year ^32^. All these facts prompted us to develop a smartphone application called Nose-Keeper which utilize artificial intelligence and may drive the development of the primary healthcare industry. With the support of the Internet, especially in developing countries and areas with high incidences of NPC, Nose-Keeper can be used to improve the care of nasal health and reduce medical costs. It can be predicted that with the further increase of the penetration rate of smartphone devices and the Internet in the future, the availability of Nose Keeper will be greatly improved, and it can effectively provide as many patients with fast, convenient and non-professional primary diagnosis services.

NPC is a severe public health problem in the underdeveloped Southeast Asian countries. In Indonesia, 13,000 new cases of NPC are reported every year ^33^. NPC is the fifth most common cancer and ninth most common cancer in Malaysia and Vietnam ^34,35^. Unfortunately, NPC is characterised by its high invasiveness and early metastasis. Owing to the insufficient number of experts in these countries, lack of sufficient experience among most primary care providers, and lack of medical awareness and adequate financial income among patients themselves, misdiagnosis and delayed diagnosis often occur, seriously threatening the lives of patients. Many patients with NPC are at an advanced stage when they first seek treatment ^36^. Clinically, NPC is considered to be the result of the interaction between Epstein-Barr virus (EBV) infection and genetic and environmental factors (such as drinking, smoking, and eating salted fish) ^37^. Therefore, during the actual medical consultation process, general practitioners or doctors in primary care institutions can comprehensively consider the diagnostic results, risk factors, and EB antibody results of the Nose-Keeper, thereby further improving the reliability of the diagnostic results and providing timely referrals for patients with NPC. The general public can use Nose-Keeper as a daily nasal health management tool. Specifically, the public can purchase high-resolution electronic nasal endoscopes on the market, learn about and use the endoscope under an instruction manual, and regularly upload nasal images to Nose-Keeper. In addition, our experiments show that Nose-Keeper is even more sensitive to NPC than a clinician with nine years of experience; therefore, it may also be used as an auxiliary tool to reduce the work stress of experts.

Recently, several deep learning studies on NPC have been published. In 2018, Li et al. ^38^ used 28,966 white-light endoscopic images to develop a deep learning model for detecting the normal nasopharynx, NPC, and other nasopharyngeal malignant tumours. Their model performance surpassed that of experts, with an overall accuracy of 88.7%. In 2022, Xu et al. ^39^ developed a deep learning model using 4,783 nasopharyngoscopy images to identify NPC and non-NPC (inflammation and hyperplasia). In 2023, He et al. ^40^ developed a deep learning model using 2,429 nasal endoscopy video frames and an algorithm named You Only Look Once (YOLO) for real-time detection of NPC in endoscopy videos. The sensitivity of their system for the detection of NPC was 74.3%. Compared with these studies, our work has the following highlights. First, our dataset consisted of 39,340 white-light endoscopic images containing both NPC and six categories of non-NPCs. The scale of our dataset is the largest ever, and the images were from three hospitals in areas with a high incidence of NPC, indicating that our dataset is more representative of real-world data. Second, by verifying many deep learning models with different architectures, we found that the Vision Transformers using the transfer learning strategy were better than the convolutional neural network using the transfer learning strategy for diagnosing NPC, providing model development guidance for subsequent researchers. The third and most important point is that we developed the Nose-Keeper. This is the first smartphone-based cloud application for NPC diagnosis in the world and is convenient to operate. To ensure the safety of the Nose-Keeper, we used the Grad-CAM algorithm to explain the decision-making process of the Nose-Keeper model visually and compared it with that of nine otolaryngologists. In addition, Nose-Keeper can identify five common diseases in daily life that are similar in appearance and clinical manifestations to NPC. These category settings make Nose-Keeper more reasonable and reliable. In fact, in areas with a high incidence of NPC, the number of non-NPC patients is far greater than the number of NPC patients. Therefore, Nose-Keeper can also provide convenient primary diagnostic services for numerous non-NPC patients and reduce their concerns about NPC, thereby alleviating the burden on the local medical system. Likewise, people in low-incidence areas of NPC can use Nose-Keeper as a nasal health management tool in their daily lives.

The application of the Nose-Keeper in healthcare in developing countries is expected to have a significant positive impact. Its primary advantage is that it significantly improves the early diagnosis of NPC and other diseases. Given that developing countries may lack skilled medical professionals and advanced medical facilities, the preliminary screening features of smartphone applications can significantly enhance early detection rates. This is especially important for decreasing misdiagnoses or missed diagnoses, which helps save medical resources and reduces reliance on more expensive and sophisticated treatment plans. In addition, the Nose-Keeper can be used as an educational tool to raise public knowledge of NPC and other frequent nasopharyngeal disorders, particularly in places with limited medical education resources. Primary healthcare professionals (PHCPs) play a significant role in developing countries. A Nose-Keeper is an essential auxiliary tool for accurate and effective disease diagnosis. In the future, the Nose-Keeper will incorporate advanced artificial intelligence algorithms, improve its ability to recognise a broader spectrum of nasal and ENT ailments and serve as a comprehensive diagnostic tool for various nasal health issues. Simultaneously, artificial intelligence will be employed to deliver personalised health advice based on user-specific data, such as lifestyle and environmental changes, to lower the risk of illness. By analysing anonymised aggregate data from users, Nose-Keeper can discover trends and patterns in NPC and other ENT disorders, providing helpful information for public health policies and resource allocation, particularly in areas with high disease incidence. Finally, recognising the value of education in health management, the Nose-Keeper will undertake an education campaign on NPC and general nasal health to raise public awareness and early detection rates. The adoption of these functions and goals will allow Nose-Keeper to play an increasingly crucial role in the healthcare systems of developing countries.

We acknowledge several limitations in our study. First, the absence of prospective testing due to dataset constraints may influence the confidence healthcare professionals and common users have in the Nose-Keeper. Second, Nose-Keeper currently lacks an image quality control function, which can result in unreliable outputs when processing images of substandard quality, such as those with blur, uneven lighting, or improper angles. Third, the datasets used were collected with professional equipment in medical settings. Therefore, biases inherent in our dataset, such as variations in image quality, imaging protocol, and imaging view, might limit the applicability of Nose-Keeper in non-clinical environments. There is a need to validate the performance of our model on images captured by household endoscopes, which have not yet been included in our dataset. Fourth, Nose-Keeper requires internet connection to link up with the cloud-based AI model for lesions detection. For some developing countries that have not yet popularize the Internet, Nose-Keeper’s availability is relatively limited. Therefore, there is a need to develop lightweight AI models to eliminate the need for high-performance devices in our next-generation Nose-Keeper, so as to achieve both cloud and local deployment simultaneously. Fifth, all patients in this study were Chinese. For safety reasons, it is crucial to use endoscopic data from people living in other high-risk areas (Such as Vietnam, Indonesia and Malaysia) to test Nose-Keeper. Future work will focus on prospective testing, developing an independent image quality control system, collecting images of different people using household nasal endoscopes and constructing a larger dataset including various image qualities. Additionally, recognizing that nasopharyngeal carcinoma diagnosis must also consider a patient’s clinical information like gender, age, dietary habits, and genetic factors, future studies will aim to develop multimodal deep learning models that integrate these variables.

The Transformer model has been widely applied in multimodal learning tasks and has achieved great success, becoming the backbone of multimodal models ^41^. At the same time, using multimodal clinical information for medical diagnosis has become a common practice in modern medicine ^42^. Especially, our work has fully validated that the Transformer model can effectively diagnose diseases such as nasopharyngeal carcinoma. Based on these insights, we have developed a strategic roadmap for integrating multimodal data (**Figure 7, Supplementary Note 4**) to enhance the practicality of Nose-Keeper in the future. We believe that with the enrichment and improvement of endoscopic datasets, the advancement of smartphones and deep learning technologies, Transformer-based multimodal models can be used to diagnose nasal and nasopharyngeal lesions in patients. More precisely, the future Nose-Keeper will use Transformers to extract and integrate clinical information (text input) of patients and endoscopic images (image input) from different regions of the nasal cavity (such as inferior turbinate, middle turbinate and nasopharynx), effectively detecting whether patients have one or more diseases. Especially, the fused multimodal features can be further utilized to generate image captions, which can help primary healthcare professionals and the public better understand various lesions ^43^. In addition, another key point of the future Nose-Keeper is to utilize model compression technology (Such as knowledge distillation, model pruning and model quantization) to reduce the device performance requirements of applications, thereby achieving local deployment and efficient cloud deployment.

**Figure 7:**
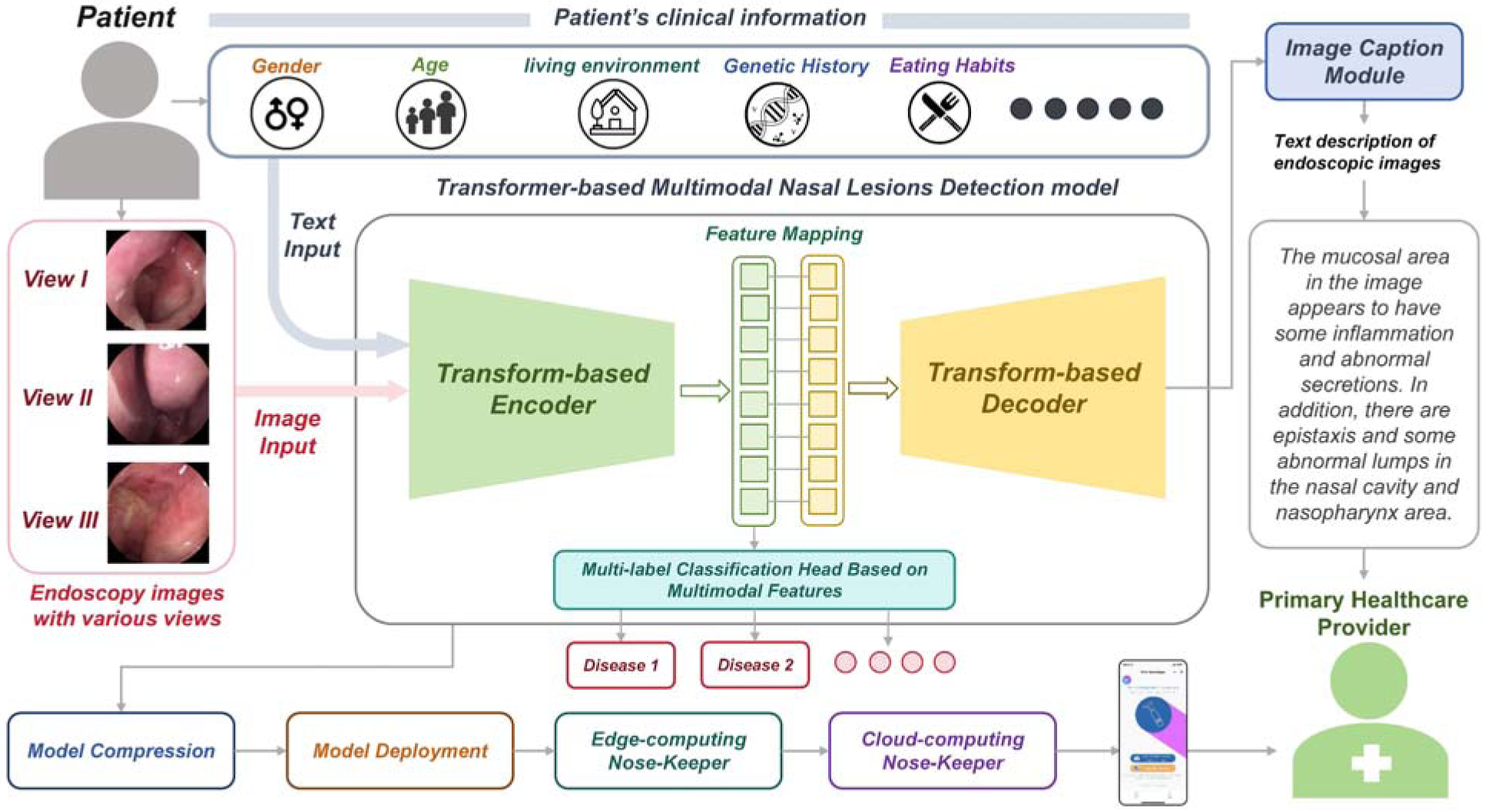
A Transformer-based strategic roadmap for enhancing the diagnostic capability and clinical utility of Nose-Keeper. On this image, the multi-modal information fusion strategy combined with clinical information in subsequent studies is shown, providing a research direction for relevant researchers.

In summary, this study represents a significant advancement in NPC diagnostics through developing Nose-Keeper, a smartphone cloud application based on cutting-edge Swin Transformer technology. Our findings demonstrate that Nose-Keeper surpasses the diagnostic sensitivity of nine professional otolaryngologists in diagnosing NPC. This was achieved by analysing a diverse and extensive nasal endoscopic dataset from multiple centres, supported by testing eight different deep learning models. Nose-Keeper’s user-friendly interface enables both medical professionals and the general public to upload endoscopic images and receive real-time AI-based preliminary screening results. Nose-Keeper is especially crucial in regions with limited access to specialized nasal endoscopy services since it not only improves primary diagnosis accuracy but also enhances awareness of NPC among primary healthcare providers and residents in high-risk areas. Furthermore, the study lays groundwork for future research into mobile healthcare and cancer detection, expanding the potential impact of DL-based smartphone app across other medical fields.

## Methods

This study was divided into three main parts: collecting datasets, constructing deep learning models, and developing mobile applications. **Figure 8** illustrated this workflow. The details of the study were comprehensively presented in the following subsections.

**Figure 8:**
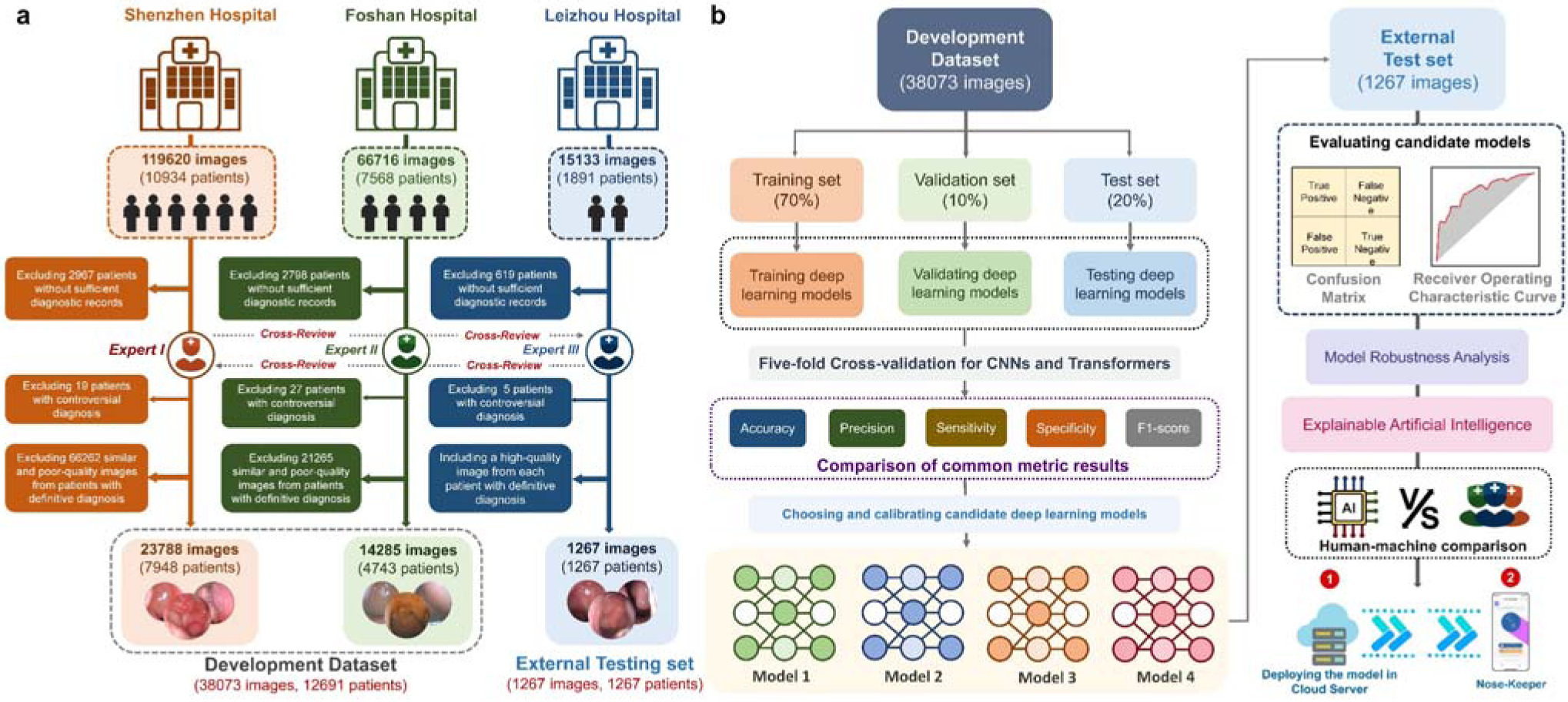
The flow chart of our work. This flowchart illustrates in detail the process of dataset construction and how to develop Nose-Keeper using deep learning models. **a** The collection process of datasets. **b** The development process of deep learning models and our smartphone application.

### Construction of the Multi-Centre Dataset

In this study, we reviewed and constructed a dataset from three hospitals located in high-risk areas of nasopharyngeal carcinoma. We retrospectively collected numerous white-light nasal endoscopic images of patients with NPC from the Department of Otolaryngology of the Second Affiliated Hospital of Shenzhen University (SZH) and the Department of Otolaryngology of Foshan Sanshui District People’s Hospital (FSH) between 1 January 2014 and 31 January 2023. Given that the early clinical symptoms of NPC (such as headache, cervical lymph node enlargement, nasal congestion, and nosebleeds) are similar to those of common diseases of the nasal cavity and nasopharynx ^44^, and rhinosinusitis, allergic rhinitis, and chronic sinusitis may be risk factors for NPC ^45–47^, we collected white-light nasal endoscopic images of non-NPC patients visiting SZH and FSH from the same period to develop deep learning models. In addition, Leizhou People’s Hospital (LZH) provided nasal endoscopic images of patients who visited the Department of Otolaryngology between 1 January 2015 and 31 April 2022. From an application perspective, including the images of non-NPC patients in the dataset can effectively improve the comprehensiveness and accuracy of the results of the deep learning model for diagnosing nasal endoscopic images. The collected images were divided into seven categories (**Figure 9**): NPC (**Figure 9a**), adenoidal hypertrophy (AH) (**Figure 9b**), allergic rhinitis (AR) (**Figure 9c**), chronic rhinosinusitis with nasal polyps (CRP) (**Figure 9d**), deviated nasal septum (DNS) (**Figure 9e**), normal nasal cavity and nasopharynx (NOR) (**Figure 9f**) and rhinosinusitis (RHI) (**Figure 9g**). **Table 4** presents the detailed characteristics of the dataset.

**Figure 9:**
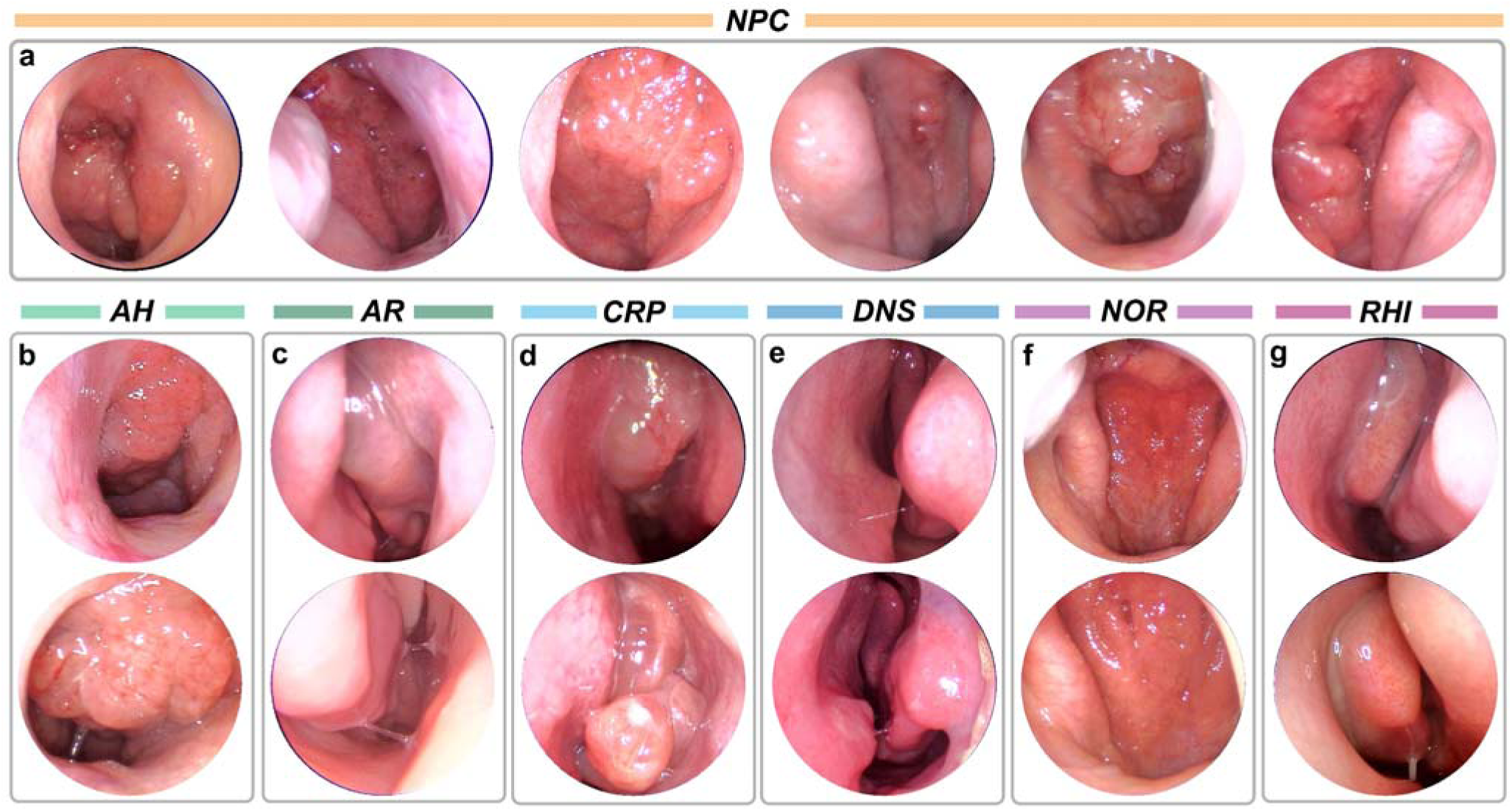
Some typical endoscopic images of different diseases. These endoscopic images are given from different angles and parts for each disease type. **a** Nasopharyngeal carcinoma (NPC). **b** Adenoidal hypertrophy (AH). **c** Allergic rhinitis (AR). **d** Chronic rhinosinusitis with nasal polyps (CRP). **e** Deviated nasal septum (DNS). **f** Normal nasal cavity and nasopharynx (NOR). **g** Rhinosinusitis (RHI).

**Table 4.** The characteristic of datasets from various hospitals, Male: M; Female: F.

| Characteristics | SZH Dataset | FSH Dataset | LZH Dataset |
| --- | --- | --- | --- |
| No. of collected images | 119,620 | 56,716 | 15,133 |
| No. of patients | 12,583 | 7,568 | 1,891 |
| Race of the patients | Asian | Asian | Asian |
| No. of qualified images | 23,788 | 14,285 | 1,267 |
| No. of patients with definitive diagnosis | 7,948 | 4,743 | 1,267 |
| Age range/Mean | 4-96/37 | 2-95/35 | 3-97/46 |
| Sex: n (%) | M: 4019(50.57%) | M: 2638(55.63%) | M: 591(46.65%) |
|  | F: 3929(49.43%) | F: 2105(44.37%) | F: 676(53.35%) |
| Level of hospital | Tertiary | Tertiary | Secondary |
|  | General Hospitals | General Hospitals | General Hospitals |
| Location of hospital | Shenzhen, | Foshan, | Leizhou, |
|  | Guangdong, China | Guangdong, China | Guangdong, China |
| The operating systems and endoscopes for obtaining images | System: Olympus | System: STORZ Xenon<br>Nova 300<br>Endoscope: STORZ<br>Image 1 HUB H3-Z | System: SD-HD668P |
|  | CV-170 Digital |  | (Shenda endoscope) |
|  | Endo-Vision |  | Endoscope: Shenda, |
|  | Endoscope: Matrix<br>E2, XION GmbH |  | J0200G |
| Image resolution(pixel) | ~ 850×850 | ~ 720×570 | ~ 480×540 |

This study was approved by the Ethics Committee of the Second Affiliated Hospital of Shenzhen University, the Institutional Review Board of Leizhou People’s Hospital and the Ethics Committee of Foshan Sanshui District People’s Hospital (reference numbers: ‘BY-EC-SOP-006-01.0-A01’, ‘BYL20220531’ and SRY-KY-2023045’) and adhered to the principles of the Declaration of Helsinki. Due to the retrospective nature of the study and the use of unidentified data, the Institutional Review Boards of SZH, FSH and LZH exempted informed consent. **Supplementary Note 5** presents more detailed ethics declarations and procedures.

### Diagnostic criteria of the nasal endoscopic images

In this study, to ensure the accuracy of the endoscopic image labels, three otolaryngologists with over 15 years of clinical experience set the diagnostic criteria based on practical clinical diagnostic processes and reference literature. Specifically, the expert combined each patient’s endoscopic examination results with the corresponding medical history, record of clinical manifestations, computed tomography results, allergen testing results (such as skin-prick testing and serum allergen-specific IgE testing), lateral cephalograms, histopathological examination results, and laboratory test results (such as nasal smear examination) to further review and confirm the diagnostic results of the existing nasal endoscopic images of each patient. A diagnosis based on the aforementioned medical records was considered the reference standard for this study. Our otolaryngologists independently reviewed all data in detail before any analysis and validated that each endoscopic image was correctly matched to a specific patient. Patients with insufficient diagnostic medical records were excluded. During the review process, when an expert doubted the diagnostic results of a particular patient, the three experts jointly made decisions on the patient’s medical records and various examination results to determine whether to include the patient in this study. The standard diagnosis for seven types of nasal endoscopic images in the dataset was as follows: (1) Nasopharyngeal carcinoma: providing the standard diagnostic label for patient images directly based on histopathological examination results ^48,49^; (2) Rhinosinusitis: further combining the patient’s medical history, clinical manifestations, and computed tomography examination ^50^; (3) Chronic rhinosinusitis with nasal polyps: further combining the patient’s medical history, clinical manifestations, computed tomography results, and pathological tissue biopsy results ^51,52^; (4) Allergic rhinitis: further combining the patient’s medical history, clinical manifestations, and allergen testing or laboratory methods ^53–55^; (5) Deviation of nasal septum: further combine the patient’s medical history and clinical manifestations and secondary analyse and evaluate the shape of the nasal septum ^56^. (6) Adenoid hypertrophy: further combine the patient’s medical history, clinical manifestations, or lateral cephalograms ^57,58^. (7) Normal nasal cavity and nasopharynx: further combination of the patient’s medical history and clinical manifestations. The nasal mucosa of a normal nasal cavity should be light red, and its surface should be smooth, moist, and glossy. The nasal cavity and nasopharyngeal mucosa show no congestion, edema, dryness, ulcers, bleeding, vasodilation, neovascularization, or purulent secretions. **Table 5** details the distribution of image categories across hospitals.

**Table 5.** The description of all nasal endoscopic white light images used in this study. ‘N’ means the number of patients corresponding to the images.

| Disease Categories | Number of images (SZH) | Number of images (FSH) | Number of images (LZH) | Number of total images (N, %) |
| --- | --- | --- | --- | --- |
| NPC | 3,526 images<br>(N=1,171, 8.39%) | 2,312 images<br>(N=769, 5.51%) | 194 images<br>(N=194, 1.39%) | 6,032 images<br>(N=2134, 15.29%) |
| AH | 2,468 images<br>(N=824, 5.06%) | 2,121 images<br>(N=706, 5.06%) | 153 images<br>(N=153, 1.10%) | 4,742 images<br>(N=1683, 12.06%) |
| AR | 2,079 images<br>(N=698, 5.00%) | 1,288 images<br>(N=424, 3.04%) | 112 images<br>(N=112, 0.80%) | 3,479 images<br>(N=1234, 8.84%) |
| CRP | 3,124 images<br>(N=1,044, 7.48%) | 797 images<br>(N=261, 1.87%) | 130 images<br>(N=130, 0.93%) | 4,051 images<br>(N=1435, 10.28%) |
| DNS | 4,316 images<br>(N=1446, 10.36%) | 2,685 images<br>(N=892, 6.39%) | 233 images<br>(N=223, 1.60%) | 7,234 images<br>(N=1435, 18.42%) |
| NOR | 5,931 images<br>(N=1,981, 14.19%) | 4,491 images<br>(N=1,495, 10.71%) | 347 images<br>(N=347, 2.49%) | 10,769 images<br>(N=3823, 27.39%) |
| RHI | 2,344 images<br>(N=784, 5.62%) | 591 images<br>(N=196, 1.40%) | 98 images<br>(N=98, 0.70%) | 3,033 images<br>(N=1078, 7.72%) |
| Total | 23,788 images | 14,285 images | 1,267 images | 39,340 images |
| Images | (N=7,948, 56.94%) | (N=4,743, 33.98%) | (N=1,267, 9.08%) | (N=13,958, 100%) |

### Deep transfer learning models

Transfer learning (TL) aims to improve the performance of new tasks by leveraging pre-learned knowledge of similar tasks. It has significantly contributed to medical image analysis, as it overcomes the data scarcity problem and saves time and hardware resources ^59^.

In this study, we effectively combined deep learning models, which are popular in artificial intelligence, with this powerful strategy. To build an optimal nasopharyngeal cancer diagnostic model, we studied Vision Transformers (ViTs), convolutional neural networks (CNNs), and hybrid models based on the latest advances in deep learning in the field of computer vision. Among them were (1) Transformers: Swin Transformer (SwinT) ^25^, Multi-Axis Vision Transformer (MaxViT) ^60^, and Class Attention in Image Transformers (CaiT) ^61^. These models were selected for their ability to model long-range dependencies and their adaptability to various image resolutions, which are crucial for medical image analysis. These represent the latest shifts in deep learning from convolutional to attention-based mechanisms, providing a fresh perspective on feature extraction. (2) CNNs: ResNet ^62^, DenseNet^63^, and Xception^64^. CNNs have gradually become the mainstream algorithm for image classification since 2012, and have shown very competitive performance in medical image analysis tasks ^65^. ResNet and DenseNet set benchmarks in terms of depth and feature propagation, whereas depthwise separable convolutions of Xception offer a balance between parameter efficiency and feature extraction. (3) Hybrid Models: PoolFormer (PoolF) ^66^ and ConvNeXt ^67^. These models combine the strengths of CNNs and Transformers. They were included because of their potential to leverage both the local feature extraction of CNNs and the global context modelling of Transformers, which is advantageous for the heterogeneous nature of medical images. We then initialised the eight architectures using pretrained weights obtained by classifying the large natural image dataset ImageNet ^68^. Because the original number of nodes of the classifiers of these networks was 1000, we reset the number of nodes of their classifiers to seven to fit our dataset. After completing initialization and adjusting the classifier, we did not choose to fine-tune some layers but instead performed comprehensive training on the entire model from scratch. Moreover, we performed probability thresholding based on Softmax.

### Explainable artificial intelligence in medical image

In medical imaging, Explainable Artificial Intelligence (XAI) is critical because it fosters trust and understanding among medical practitioners and facilitates accurate diagnosis and treatment by elucidating the rationale behind AI-driven image analysis. In this study, we used Gradient-weighted Class Activation Mapping (Grad-CAM) ^69^ to generate a corresponding heatmap. Red indicates high relevance, yellow indicates medium relevance, and blue indicates low relevance. Grad-CAM helps to visualise the regions of an image that are important for a particular classification. This is crucial in medical image classification, as it helps people understand which parts of the image contribute to model decision-making and validates whether the model focuses on disease-related features. By providing visual explanations through heat maps, Grad-CAM can help build trust among medical practitioners and the public regarding the decisions made by AI systems.

### Development process of models and smartphone applications

All the nasal endoscopic images were divided into two parts. The first part contained 38073 images from SZH and FSH, which were used as the development dataset for training and validating the performance of the model. The development dataset was further divided into three parts in a 7:1:2 ratio, i.e., internal training, internal validation, and internal test sets. The second part contained 1267 images from the LZH, which were used as an external test set to test the performance of the model in real-world settings and verify the robustness of the model.

Before training the various networks, we resized all images to 224×224×3. Subsequently, the images were normalised and standardised using the mean [0.2394,0.2421,0.2381] and standard deviation [0.1849, 0.28, 0.2698] of the three channels. To improve the robustness and generalisation ability of the models, we utilised the Transformers Library provided by Pytorch to automatically transform (***RandomRotation, RandomAffine, GaussianBlur and Color Jitter***) the image inputs during training. The loss functions of All models uniformly used the cross-entropy loss function. During the training process, we employed the AdamW optimiser with a 0.001 initial learning rate, β1 of 0.9, β2 of 0.999, and weight-decay of 0.0001 to optimise eight models’ parameters. We set the number of epochs to 150 and used a batch size of 64 for each model training.

We adopted an early stopping strategy, which meant that the model training will be stopped automatically stopped when its accuracy (patience and min_delta were set to 10 and 0.001, respectively) on the internal validation set no longer significantly improved for some time, thereby preventing overfitting. We calibrated each model using an internal validation set with temperature scaling, a method for calibrating deep learning models and assessed the calibration performance using the Brier-Score and Log-Loss. During the validation and inference stages, the model’s image preprocessing process was consistent with the training stage, but automatic image transformation was no longer implemented. We used the PyTorch framework (version 2.1), a computer with the Ubuntu 20.04 system and an NVIDIA GeForce RTX 4090 to complete the entire experiment. The weights of all models were saved in ‘Pth’ format.

In this study, we developed a responsive and user-friendly Android application that prioritises maintainability and scalability. Utilising Java for native Android development, we embraced the MVVM design pattern for application modularisation, incorporating bidirectional data binding for seamless UI and data synchronisation. Our tech stack included Retrofit for network requests alongside third-party libraries like ButterKnife, Gson, Glide, EventBus, and MPAndroidChart for enhanced functionality and user experience, complemented by custom animations and NDK for hardware interaction. At the backend, we leveraged SSM (Spring + SpringMVC + MyBatis), Nginx, and MySQL for a high-performance architecture. For database, we used MySQL to manage data and adopted Redis for caching. The backend of the application and deep learning model were deployed on a high-performance Cloud Server (Manufacturer: Tencent; Equipment Type: Standard Type S6; Operating System: Centos 7.6; CPU: Intel® Xeon® Ice Lake; Memory: DDR4) with Nginx load balancing to optimise server resource utilisation (See **Supplementary Note 1** for details). To ensure the security of applications and personal privacy data, we used encryption protocols and algorithms and toolkits that comply with industry standards (See **Supplementary Note 2** for details). When utilizing Nose-Keeper, all input images must go through an image preprocessing pipeline consistent with the model inference stage.

### Model evaluation and statistical analysis

For the development datasets (SZH and FSH), eight models were evaluated using five standard metrics: overall accuracy (equation (1)), precision (equation (2)), sensitivity (equation (3)), specificity (equation (4)), and f1-score (equation (5)). The definitions of these five metrics were as follows (See **Supplementary Note 3** for details).

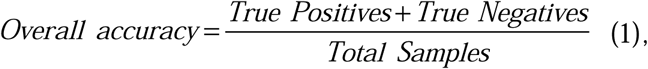

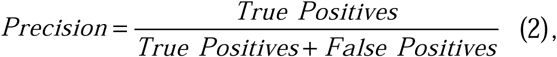

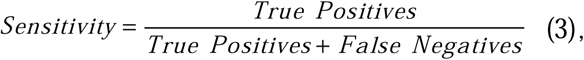

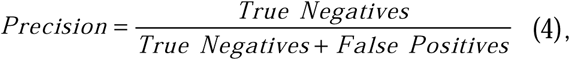

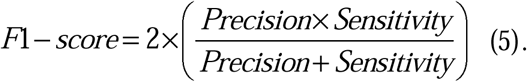

To avoid performance uncertainty caused by random splitting of the development dataset, we used a five-fold cross-validation strategy to evaluate the potential of various models on the development dataset and then selected four more excellent models from the eight models that can be used for the smartphone application based on the quality of the metric results. After selecting the four candidate models, we used a confusion matrix and Receiver Operating Characteristic (ROC) curve to further evaluate the performance of the candidate models in an external test set (LZH). A larger area under the ROC curve (AUC) indicated better performance. We used the Beset model to develop a smartphone application. Statistical analyses were performed using Python 3.9. Owing to the large sample size of the internal dataset and the use of five-fold cross-validation, we used the normal approximation to calculate the 95% confidence intervals (CI) of overall accuracy, precision, sensitivity, specificity, and f1-score. In the external test set, we used an Empirical Bootstrap with 1000 replicates to calculate the 95% CI of the AUC. The 95% CIs of overall accuracy, sensitivity and specificity were calculated using the Wilson Score approach in the Statsmodels package (version 0.14.0).

### Analysing the Robustness of the deep learning models via data augmentation

The use of images with different data augmentations to test the model can reveal its adaptability to input changes and analyse its robustness ^70^. In particular, data augmentation simulates possible image transformations in practical applications, thereby testing the stability and performance of a model when faced with unseen or changing images. This strategy helps developers identify the potential weaknesses of the model, guide subsequent improvements, and enhance the application reliability of the model in complex and ever-changing environments. We used an external test set to analyse the prediction result changes of the model under ***Gaussian blur***, ***Saturation changes***, ***Image rotation*** and ***Brightness changes***. Prior to testing the model, we augmented the external test set using Pillow (version 9.3.0). For each transformation, we assigned different parameter values to the built-in functions of Pillow, resulting in 12 enhanced datasets from the external datasets.

### Comparison of the diagnostic performance between deep learning model and clinicians

The representativeness of the external test set is crucial for fully comparing the performance differences between AI and human experts. Therefore, when retrospectively collecting endoscopic images, in addition to ensuring the accuracy of image labels, our expert team also fully considered the severity of lesions, different stages of disease, and differences in appearance in each endoscopic image. Meanwhile, the expert group also ensured as much as possible the age difference and gender balance of the entire dataset. Especially, the time span of external test set has reached five years. We recruited nine otolaryngologists with different clinical experiences from three institutions, i.e., one year (two otolaryngologists), three years (one otolaryngologist), four years (one otolaryngologist), five years (one otolaryngologist), six years (one otolaryngologist), eight years (two otolaryngologists), and nine years (one otolaryngologist). Before each expert independently evaluated the external test set, we shuffled dataset and renamed each image as “test_xxxx. jpg” and distributed it to all experts. We required experts to independently evaluate each endoscope within a specified time frame to simulate the physical and mental stress faced by experts in actual clinical settings, which further reflects the efficiency of AI. Notably, we prohibited experts from consulting diagnostic guidelines and mutual communication. All expert evaluation results were anonymized and automatically verified through a python program. Finally, we plotted a diagnostic performance heatmap, confusion matrix, ROC curve, and optimal Youden-index to comprehensively and intuitively demonstrate the performance differences between AI and clinicians in diagnosing different diseases.

## Data availability

A subsample of the internal test set with 110 images per diagnostic class is available upon reasonable request from the authors.

## Code availability

The custom codes and model weights for Nose-Keeper development and evaluation in this study are accessible on GitHub (https://github.com/YubiaoYue/Nose-Keeper).

## Acknowledgments

The author is supported by the NSF of Guangdong Province (No.2022A1515011044, No.2023A1515010885), and the project of promoting research capabilities for key constructed disciplines in Guangdong Province (No.2021ZDJS028).

## Author contributions

Conception and design: Y.Y., X.Z., H.L. and Z.L. Funding obtainment: Z.L. Provision of study data: X.Z., F.Z., K.Z. and L.L. Collection and assembly of data: X.Z., F.Z., J.X. and L.L. Data analysis and interpretation: Y.Y., X.Z. and Z.L. Manuscript writing: Y.Y., X.Z., H.L. and Z.L. All authors have read and approved the manuscript.

