## Supplementary Information for "A Deep Learning Based Smartphone Application for Early Detection of Nasopharyngeal Carcinoma Using Endoscopic Images"

### Contents


| **Supplementary Item** | **Page** |
|---|---|
| Supplementary Figure 1 | 3 |
| Supplementary Figure 1 | 4 |
| Supplementary Table 1 | 5 |
| Supplementary Note 1 | 6 |
| Supplementary Note 2 | 7 |
| Supplementary Note 3 | 9 |
| Supplementary Note 4 | 11 |
| Supplementary Note 5 | 12 |
| Supplementary Note 6 | 13 |
| Supplementary Movie 1 | 14 |

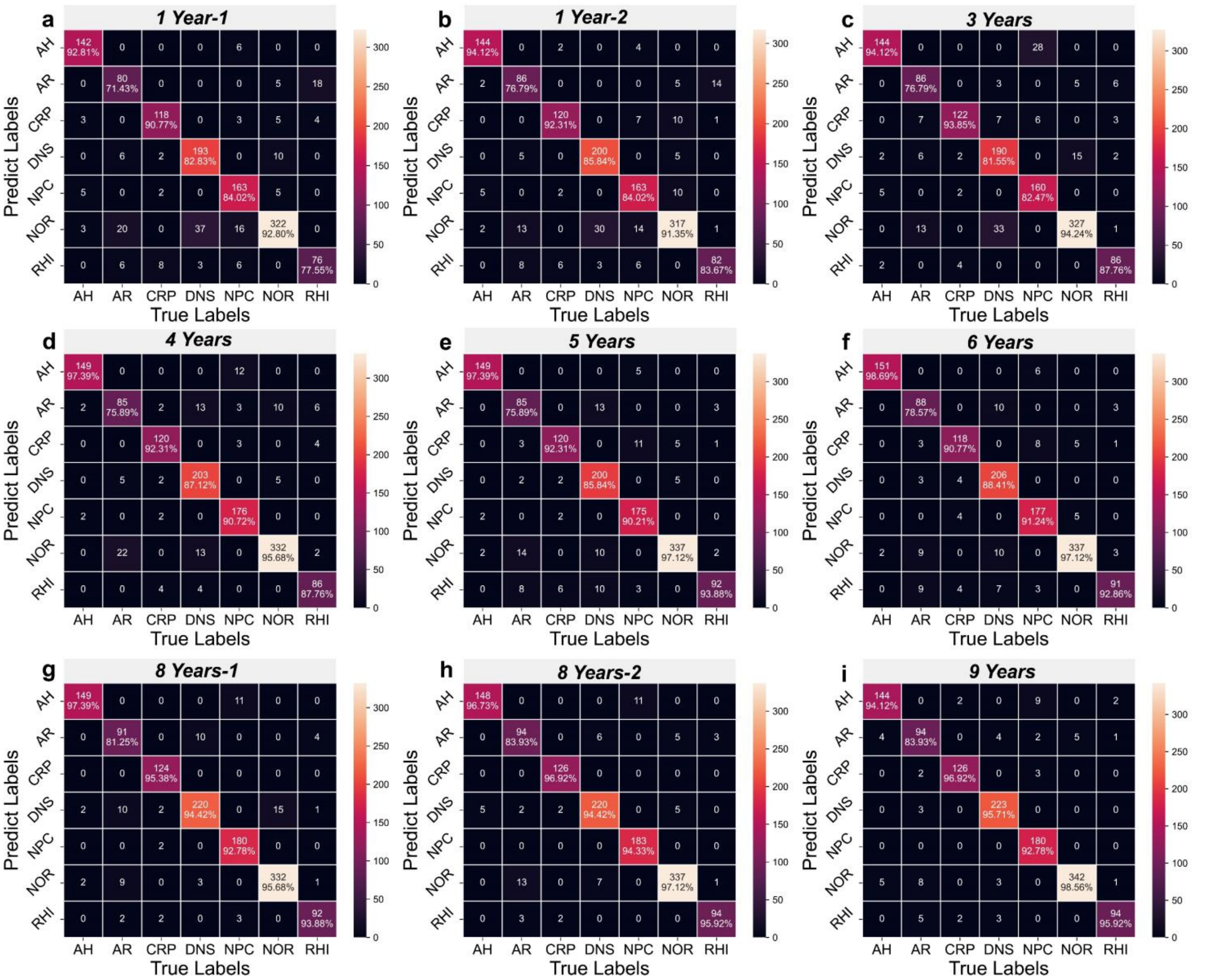


**Supplementary Figure 1. The confusion matrix of nine clinicians**. A total of 9 otolaryngologists participated in the human-computer comparison experiment. **a** The confusion matrix of a clinician with one year of clinical experience. **b** The confusion matrix of a clinician with one year of clinical experience. **c** The confusion matrix of a clinician with three years of clinical experience. **d** The confusion matrix of a clinician with four years of clinical experience. **e** The confusion matrix of a clinician with five years of clinical experience. **f** The confusion matrix of a clinician with six years of clinical experience. **g** The confusion matrix of a clinician with eight years of clinical experience. **h** The confusion matrix of a clinician with eight years of clinical experience. **i** The confusion matrix of a clinician with nine years of clinical experience.

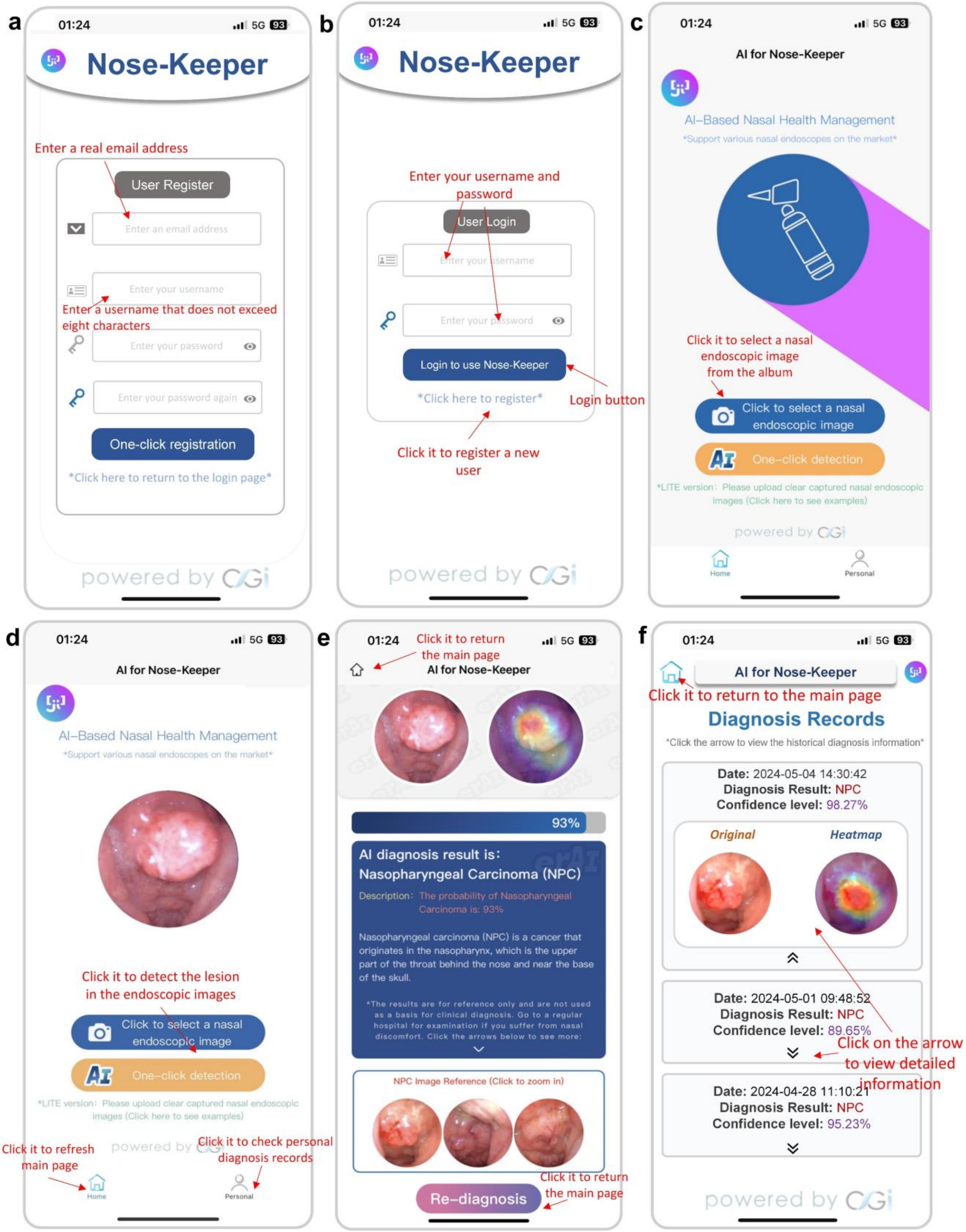


**Supplementary Figure 2. An introduction to Nose-Keeper's user interface and usability features.** We have used red arrows in the figure to illustrate the functions of each button of Nose-Keeper. **a** The registration page. **b** The login page. **c** The home page (data upload). **d** The page after uploading the image. **e** The result page after data processing. **f** The diagnostic records page.

**Supplementary Table 1.** The time required for performing the experiments.

| Model | Train time/ One-epoch (s) | Val time/ One-epoch (s) | Test time/ One-epoch (s) | All time for five-fold validation | External test / One-epoch (s) |
|---|---|---|---|---|---|
| Xception | 70.5-73.3 | 4.5-4.8 | 12.5-13.2 | 7 hours,38 minutes,44 seconds | 3.8 |
| SwinT | 49.5-54.2 | 4.3-4.5 | 12.8-13.0 | 5 hours,8 minutes,51 seconds | 3.6 |
| CaiT | 105.3-107.6 | 6.2-6.3 | 14.4-15.1 | 6 hours,15 minutes,33 seconds | N/A |
| PoolFormer | 78.9-81.8 | 4.8-5.0 | 13.7-13.9 | 7 hours,43 minutes,19 seconds | 4.1 |
| ConvNeXt | 135.8-141.5 | 5.8-6.0 | 14.6-14.9 | 8 hours,21 minutes,25 seconds | 4.6 |
| DenseNet | 92.1-95.4 | 4.9-5.2 | 13.7-14.0 | 7 hours,30 minutes,38 seconds | N/A |
| ResNet | 61.2-63.8 | 4.1-4.4 | 13.4-13.6 | 5 hours,13 minutes,54 seconds | N/A |
| MaxVit | 266.1-271.9 | 13.6-14.2 | 26.2-26.9 | 23 hours,49 minutes,53 seconds | N/A |

**Supplementary Note 1: Development Details of the Smartphone Application**

Our goal is to develop a responsive, user-friendly, and efficient smartphone application (based on the Android platform) while ensuring the maintainability and scalability of our codes. Throughout the development process, we adopted modern software engineering methods and technology stacks.

We used native Android development based on Java language for front-end development and adopted a design pattern based on Model-View-ViewModel (MVVM) to promote modularization of applications. In our architecture, the View layer is responsible for displaying the user interface, the ViewModel layer handles business logic and data operations, and the Model layer manages data. By bidirectional data binding between ViewModel and View, the interface and data are effectively synchronized and updated. In terms of network requests, we have adopted the Retrofit library to package the API interfaces to improve the efficiency and maintainability of network requests. Besides, our application also integrates various third-party libraries, including ButterKnife to simplify view binding and event handling; Gson is used for serialization and deserialization of JSON data; Glide is used for efficiently loading images; EventBus reduces the coupling between components; MPAndroidChart implements complex data visualization requirements. Meanwhile, we also implemented specific animation effects through custom views in the application to optimize the user experience. The calling and image processing of related hardware devices are achieved through NDK, ensuring the performance and stability of the application.

The back-end development adopted technologies such as SSM (Spring + SpringMVC + MyBatis), Nginx, and MySQL, forming a high-performance front-end and back-end separation architecture. We follow the MVC architecture pattern to improve the manageability and maintainability of our code. At the database level, we use a combination of MySQL and Redis. MySQL is used to process structured data, while Redis is a key value pair database used for high-speed caching. The combination of the two maximizes the efficiency of data storage and retrieval. The entire application and platform are deployed on Tencent Cloud Server (Standard Type S6; Operating System: Centos 7.6; CPU: Intel) ® Xeon ® Ice Lake; Memory: DDR4), and optimized the deployment of web applications using Nginx's load balancing and proxy technology, thereby improving the efficiency of server resource utilization.

**Supplementary Note 2: Security Policy for Nose-Keeper**

**(1) Software packages or tools for data transfer with SSL/TLS encryption.** Nose-Keeper uses HTTPS protocol (TLS) to encrypt the communication between the front-end and back-end. We applied for the TrustAsia server certificate, used the HttpsURLConnection client library in the client, and configured SSLSocketFactory and TrustManager to handle SSL/TLS certificates. We configured the client using Fetch, which supports the HTTPS protocol. On the server side, we used Spring Boot to configure Server.ssl to specify the location and password of the SSL/TLS certificate file. Spring Security was used to implement authentication and authorization based on OAuth2.0.

**(2) The exact industry-standard encryption protocols and algorithms used for encrypting sensitive user data.** Nose-Keeper's sensitive data mainly includes user registration information (email address, login password), original images collected by users, and analyzed heat map information. In order to meet the confidentiality requirements of different data, Nose-Keeper mainly uses AES-256 and SHA-256 algorithms. In order to ensure that user passwords are not monitored and intercepted when logging in, we use SHA-256 to convert registration information into a fixed-length hash value and store it in the server. SHA-256 is widely regarded as one of the most secure hash functions today and can be used to improve the security of personal information and password storage. In addition, its output length is only 256 bits and its calculation speed is relatively fast. For image-type sensitive data, we use AES-256 for encryption and decryption. AES-256 is a symmetric encryption algorithm that uses block encryption with a key length of 256 bits. Due to its high security and reliability and the acceleration support of modern hardware for this algorithm, AES-256 shows extremely high efficiency when processing large-scale data. These characteristics make AES-256 particularly suitable for encrypting and decrypting large-capacity data such as images. In addition, we also protect all data exchange processes between the front-end and back-end through the HTTPS protocol to ensure communication security.

**(3) Data Protection Regulations.** In terms of the data protection regulations, the Nose-Keeper complies with the General Data Protection Regulation and the Regulations on Management of Mobile Internet Application Information Service.

**(4) Protocols for Accessing or decrypting sensitive personal data.** Nose-Keeper uses SHA-256 to encrypt the user's login information and transmits it securely to the back-end using the HTTPS protocol. After obtaining the encrypted user login information, the back-end first decrypts the information using the HTTPS protocol and then uses SHA-256 to encrypt the user information again and store it in the database. Note that since SHA-256 is a one-way encryption algorithm, the user's personal information cannot be decrypted under any circumstances (only the user knows the original data). When a user uses Nose-keeper, Nose-Keeper will use AES-256 by default to decrypt the images collected by the user and the analyzed heat map and display them on the corresponding front-end page. Each user has the right to obtain, modify, and delete any of their own data (images collected by each user, analyzed images, and his registration information). Unauthorized personnel/other users cannot access or decrypt any data that does not belong to them.

**Supplementary Note 3: Metrics used for model evaluation**

(1) Overall accuracy

Overall accuracy is the most intuitive performance metric, representing the proportion of correctly classified samples in the total sample size. In nasopharyngeal carcinoma diagnosis, high accuracy means that the model can effectively distinguish between NPC and non-NPC diseases, which is crucial for evaluating the overall performance of the model.

(2) Precision

Precision refers to the proportion of samples that are true positive (e.g., nasopharyngeal carcinoma) among all samples predicted by the model to be positive. In medical image classification, high precision reduces the likelihood of misdiagnosis and is crucial for ensuring the accuracy of treatment and intervention measures.

(3) Sensitivity

The sensitivity measure models the ability to correctly identify true positive samples. In nasopharyngeal carcinoma detection, high sensitivity means that the model can effectively identify the majority of true cases of the disease, which is crucial for early detection and treatment.

(4) Specificity

Specificity measures the model's ability to correctly identify negative samples (e.g., non-NPC lesions). High specificity reduces false positive rates and is particularly important to avoid unnecessary further testing and treatment.

(5) F1-Score

The F1-score is a harmonic mean of precision and recall, and is a comprehensive performance metric. Because it is often necessary to balance recall and precision in medical diagnosis, the F1 score provides a method for quantifying this balance.

(6) Confusion Matrix

The confusion matrix provides an intuitive way to view the performance of the model on various categories, including true positives, false positives, true negatives, and false negatives. It helps reveal the bias of the model on specific categories and is crucial for refining model performance and guiding subsequent improvement efforts.

(7) Receiver Operating Characteristic Curve (ROC Curve) and AUC

The ROC curve provides a method for evaluating the discrimination ability of a model by plotting the true positive rate (sensitivity) and false positive rate (1-specificity) of the model at different thresholds. The area under the curve (AUC) quantifies the average performance of the model across all possible classification thresholds, with a higher AUC value indicating better discrimination ability.

In summary, these metrics provide a comprehensive framework for evaluating the performance of deep learning models on classification tasks such as nasopharyngeal cancer and other common lesions. They not only help assess the overall effectiveness of the model, but also enable researchers to deeply analyze the strengths and weaknesses of the model, guiding further optimization and improvement of the model.

**Supplementary Note 4: Subsequent improvements to the model**

(1) **Expanding the Dataset.** To ensure that our findings are generalizable, we intend to test the model on a more extensive and diverse dataset, which includes images from a broader range of patient demographics and a larger variety of imaging conditions.

(2) **Interpretability and Decision Support**. Beyond accuracy, we aim to understand how the AI model makes decisions compared to human experts. By utilizing techniques such as saliency maps, SHapley Additive exPlanations and layer-wise relevance propagation, we can provide in-depth insights into the AI's decision-making process, making it a more valuable decision-support tool for clinicians.

(3) **Multi-Modal Learning**. Future improvements of the model will incorporate multi-modal data, such as medical history, clinical report, endoscopic images of different nasal locations and other diagnostic tests, to perform a more holistic and accurate analysis similar to the practice of human otolaryngologists.

**(4) Prospective testing in a clinical setting.** Collaborating with medical institutions and testing AI diagnostic tools in actual clinical settings can ensure their effectiveness and reliability in the real world. Importantly, this helps increase the confidence of patients and clinicians in the efficiency of AI, which is a necessary step in accelerating the clinical translation of AI tools.

**Supplementary Note 5: Ethical declarations, safeguards and procedures**

(1) **Ethical Approvals**. This study was approved by the Ethics Committee of the Second Affiliated Hospital of Shenzhen University, the Institutional Review Board of Leizhou People's Hospital and the Ethics Committee of Foshan Sanshui District People's Hospital (reference numbers: ‘BY-EC-SOP-006-01.0-A01’, ‘BYL20220531’ and SRY-KY-2023045’) and adhered to the principles of the Declaration of Helsinki.

(2) **Data De-identification**. Prior to the commencement of the study, all patient data were de-identified, stripping away any personal identifiers that could link the data back to individual patients. This process is strictly implemented to ensure compliance with privacy laws and that the patient's personal privacy information is not disclosed.

(3) **Exempting Informed Consent**. The Institutional Review Boards of SZH, FSH and LZH exempted informed consent. The specific reasons are as follows: the retrospective nature of the data acquisition, the use of deidentified images, the minimal risk to patients since the research does not affect clinical care, and the public interest in developing advanced diagnostic tools through AI.

(4) **Data Security Protocols**. We have implemented data security measures to safeguard the anonymized datasets. Specifically, access to data is strictly controlled and monitored, all data is stored on securely encrypted servers, and access rights to data are restricted.

(5) **Periodic review of data access**. Periodic review is conducted to ensure that data access is limited to authorized personnel and that the purpose of use is consistent with the approved research purpose. These reviews help prevent unauthorized access and ensure that all data are used in accordance with research purposes and ethical requirements.

**Supplementary Note 6: The multi-image voting mechanism of Nose-Keeper**

We recognized the critical importance of data quality and diversity in enhancing the performance of deep learning models. To address this, we implemented a robust data collection strategy that involves acquiring images from different parts of the patient's nasal cavity and imaging conditions. This approach ensures a more representative dataset, enabling the model to learn generalized features and thereby reducing misclassification rates that may arise from sensitivity to specific conditions. To further bolster diagnostic accuracy, we incorporated a multi-image voting mechanism within our system. This mechanism allows Nose-Keeper to read three consecutive endoscopic images of the lesion area from each patient and submit them for predictive analysis. A final diagnosis is only returned when at least two of the three predictions are consistent; in cases of inconsistency, the system alerts the user to potential image errors and advises retesting. This method mitigates the risk of misdiagnosis caused by issues such as poor image quality, jitter, or improper acquisition angles. Additionally, we are committed to continuous model iteration and optimization, with plans to regularly update the model with more diverse datasets and fine-tune it to adapt to evolving medical image characteristics. This iterative process is designed to ensure that our model remains accurate and reliable over time.

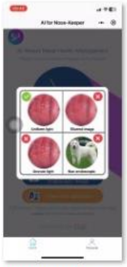

Supplementary Movie 1.mp4

**Supplementary Movie 1. A short video showing the Nose-Keeper.** We briefly demonstrated how to use Nose-Keeper to load a single local image and the Nose-Keeper results page.
